# Profile of SARS-CoV-2-specific CD4 T cell response: Relationship with disease severity and impact of HIV-1 and active *Mycobacterium tuberculosis* co-infection

**DOI:** 10.1101/2021.02.16.21251838

**Authors:** Catherine Riou, Elsa du Bruyn, Cari Stek, Remy Daroowala, Rene T. Goliath, Fatima Abrahams, Qonita Said-Hartley, Brian W. Allwood, Marvin Hsiao, Katalin A. Wilkinson, Cecilia S. Lindestam Arlehamn, Alessandro Sette, Sean Wasserman, Robert J. Wilkinson, on behalf of the HIATUS consortium

## Abstract

T cells are involved in control of COVID-19, but limited knowledge is available on the relationship between antigen-specific T cell response and disease severity. Here, we assessed the magnitude, function and phenotype of SARS-CoV-2-specific CD4 T cells in 95 hospitalized COVID-19 patients (38 of them being HIV-1 and/or tuberculosis (TB) co-infected) and 38 non-COVID-19 patients, using flow cytometry. We showed that SARS-CoV-2-specific CD4 T cell attributes, rather than magnitude, associates with disease severity, with severe disease being characterized by poor polyfunctional potential, reduced proliferation capacity and enhanced HLA-DR expression. Moreover, HIV-1 and TB co-infection skewed the SARS-CoV-2 T cell response. HIV-1 mediated CD4 T cell depletion associated with suboptimal T cell and humoral immune responses to SARS-CoV-2; and a decrease in the polyfunctional capacity of SARS-CoV-2-specific CD4 T cells was observed in COVID-19 patients with active TB. Our results also revealed that COVID-19 patients displayed reduced frequency of Mtb-specific CD4 T cells, with possible implications for TB disease progression. There results corroborate the important role of SARS-CoV-2-specific T cells in COVID-19 pathogenesis and support the concept of altered T cell functions in patients with severe disease.

## INTRODUCTION

Coronavirus disease 2019 (COVID-19), caused by severe acute respiratory syndrome coronavirus 2 (SARS-CoV-2), emerged in December 2019 and is the cause of a devastating pandemic resulting in more than 100 million infections and over 2 million deaths within the last year. COVID-19 shows an extremely variable clinical course: ranging from an asymptomatic state or mild respiratory symptoms to severe viral pneumonia with or without acute respiratory distress syndrome (Huang et al., 2020). While most COVID-19 cases are mild (∼80%), up to a quarter of SARS-CoV-2 infected persons present with severe disease necessitating hospitalization and ∼5% of critical cases require intensive care, putting extreme pressure on health systems. Severe disease is most commonly observed in those with advanced age, male gender and pre-existing co-morbidities (such as hypertension, type 2 diabetes, obesity or chronic lung disease) (Clark et al., 2020). Immunologically, COVID-19 severity has been associated with major systemic alterations of the host immune system, including profound lymphopenia, skewed distribution and activation of T cell subpopulations, disruption of the B cell compartment and elevated plasma concentrations of proinflammatory cytokines (Chen and John Wherry, 2020; Laing et al., 2020; Mathew et al., 2020; Zheng et al., 2020; Zhou et al., 2020). A growing body of evidence suggests that SARS-CoV-2-specific T cell response plays a key role in modulating COVID-19 pathogenesis (Meckiff et al., 2020; Oja et al., 2020; Rydyznski Moderbacher et al., 2020; Sattler et al., 2020; Schub et al., 2020; Weiskopf et al., 2020). And while the precise nature of T cell responses conferring protection is still unclear, it is now well established that SARS-CoV-2 elicits a broad T cell response in the majority of patients, with CD4 responses being dominant over CD8 (Altmann and Boyton, 2020). Moreover, pre-existing SARS-CoV-2 cross-reactive T cells may also contribute to the divergent manifestations of COVID-19. These cells, likely acquired during previous infections with endemic human coronaviruses, have been identified in 20 to 50% of SARS-CoV-2-unexposed individuals in different populations around the world (Braun et al., 2020; Grifoni et al., 2020; Le Bert et al., 2020; Mateus et al., 2020; Rydyznski Moderbacher et al., 2020; Sekine et al., 2020; Sette and Crotty, 2020; Weiskopf et al., 2020). It is yet to be determined whether pre-existing immunity to SARS-CoV-2 is sufficient to confer protection or attenuate the severity of COVID-19. Our knowledge regarding SARS-CoV-2 induced immune responses is rapidly expanding, yet very few studies have simultaneously examined the magnitude, functional and phenotypic profile of SARS-CoV-2-responding T cells in relation to disease severity. This represents an important gap in our understanding of the role played by T cells during the clinical course of COVID-19, which has implications for pathogenesis and the assessment of vaccine efficacy.

Importantly, in countries with a high burden of HIV-1 and *Mycobacterium tuberculosis* (Mtb) infections, the intersecting coronavirus, HIV-1 and TB epidemics pose additional public health challenges. HIV infection induces a profound dysregulation of both the innate and adaptive immune systems (Deeks et al., 2015), weakening the host’s ability to mount and/or maintain immune responses to other pathogens or upon vaccination (Avelino-Silva et al., 2016; Rubin et al., 2014). Furthermore, immune dysfunctions often persist despite anti-retroviral therapy (ART) (Klatt et al., 2013). It is therefore likely that HIV-1 infection will impair the SARS-CoV-2 immune response. Likewise, co-infection with active tuberculosis (aTB) and COVID-19 is also of particular concern. Both diseases are primarily respiratory illnesses, eliciting a hyper-inflammatory state in the lung. It is thus reasonable to speculate that the hyper-inflammatory milieu induced by COVID-19 could accelerate TB disease progression and vice-versa (Gupta et al., 2020b). Moreover, profound lymphopenia and hyper-inflammation associated with COVID-19 could favor Mtb reactivation. These concerns are further underlined by several large epidemiological studies showing that, both HIV-1 and active TB independently associate with an increased risk of COVID-19-related death (Bhaskaran et al., 2021; Boulle et al., 2020; Brown et al., 2021; Chen et al., 2020a; Davies, 2020; Geretti et al., 2020; Tamuzi et al., 2020). It is thus an urgent research priority to investigate the profile of the SARS-CoV-2-specific T cells in patients co-infected with HIV-1 and/or active TB and to assess the impact of acute SARS-CoV-2 infection on the Mtb-specific memory CD4 T cell response.

In this study, focused on pathogen-specific CD4 T cell responses, our aims were to: 1) Compare the profile of pre-existing SARS-CoV-2 cross-reactive CD4 T cells and COVID-19-induced CD4 T cells, 2) Define the relationship between COVID-19 severity and the SARS-CoV-2-specific CD4 T cell response, 3) Investigate the impact of HIV and/or active TB co-infection on the SARS-CoV-2-specific CD4 T cell response and 4) Assess the effect of COVID-19 on the Mtb-specific CD4 T cell response.

## RESULTS

### Clinical characteristics of the study participants

Using a whole blood assay, we investigated the SARS-CoV-2- and Mtb-specific CD4 T cell response in hospitalized COVID-19 (n = 95) and non-COVID-19 patients (n = 38). The clinical characteristics of patients are presented in **Table 1**. COVID-19 cases were defined based on a documented positive SARS CoV-2 PCR swab result and hospitalized non-COVID-19 patients were all SARS CoV-2 PCR negative and had no detectable SARS-CoV-2 nucleocapsid-specific IgG measured in blood collected at enrolment. Samples were collected at a median of 2 days (IQR: 1-4) after admission to hospital, Cape Town (South Africa). Briefly, the median age was comparable between the two groups (52 vs 51 years) and male predominated in the COVID-19 group (57.9% vs 34.2%, *P* = 0.014). A high proportion of co-morbidities, such as hypertension (46.3%), diabetes (32.6%) and obesity (26.3%), was reported in the COVID-19 group. The non-COVID-19 controls were well-matched to the COVID-19 group in terms of prevalence of hypertension, diabetes and obesity. However, a greater proportion of non-COVID-19 controls had cardiovascular (44.7% vs 7.3%, *P* < 0.0001) and other respiratory diseases (28.9% vs 2.1%, *P* < 0.0001) compared to the COVID-19 group. COVID-19 patients had a range of different requirements for oxygen therapy and supportive care, as reflected by their World Health Organization (WHO) ordinal scale score (see methods), with approximately half being classified as mild/moderate cases (WHO < 5) and the other half as severe cases (WHO ≥ 5). Most non-COVID-19 controls did not require oxygen therapy (57.9%). The majority of the COVID-19 patients received treatment with steroids (78.9%) following the outcome of the RECOVERY trial (Recovery Collaborative Group et al., 2020). About 1/3 of the recruited participants were HIV-1 infected (n = 31). In the COVID-19 group, the majority of HIV-1-infected patients were on ART (74.2%) with a median CD4 count of 132 cells/mm^3^ and a median log viral load <1.3 log mRNA copies/ml. HIV-1-infected non-COVID controls had a lower median CD4 count (20 cells/mm^3^, *P* = 0.03) and higher viral loads (5.37 log mRNA copies/ml, *P* = 0.0005) owing to proportionally fewer participants being on ART in this group (46.1%). Lastly, 15 participants in the COVID-19 group had active TB (8 of them also being HIV-1 infected) and 5 non-COVID-19 controls had active TB (all of them being HIV-1 infected). It is important to mention that most of the HIV-1 infected participants without active TB were virally suppressed (77.3%, 17/22), while only one of the seven HIV-1 infected participants with active TB was aviremic.

**Table 1:** Clinical characteristics of COVID-19 patients and SARS-CoV-2-uninfected hospitalized controls (non-COVID). ^*^: SARS-CoV-2 serology was performed using the Roche Elecsys assay, measuring SARS-CoV-2 nucleocapsid-specific antibodies. Medians are reported and numbers in brackets correspond to interquartile range [IQR]. ART: antiretroviral treatment, COI: Cut-off index of Roche Elecsys assay, CRP: C-Reactive protein, LDH: lactate dehydrogenase.

|  | **COVID-19 (n=95)** | **NON-COVID (n=38)** | **P-values** |
| --- | --- | --- | --- |
| **Age (median, IQR)** | **52 [43-57]** | **51 [38-66]** | 0.87 |
| **Male (%)** | **57.9%** | **34.2%** | **0.014** |
| **SARS-CoV2 PCR positive** | **100%** | **0%** | - |
| **SARS-CoV2 serology positive\*** | **70.5% (n=67)** | **0%** | - |
| COI (median, IQR) | **7.06 [0.32-27.06]** | **0.07 [0.07-0.08]** | - |
| **HIV positive (% , n)** | **32.6% (n=31)** | **34.2% (n=13)** | 0.86 |
| on ART | **74.2% (n=23)** | **46.1% (n=6)** | 0.07 |
| Time on ART (years) | **9.6 [6-12]** | **3.6 [0.4-11]** | 0.22 |
| CD4 count (cells/mm<sup>3</sup>) | **132 [51-315]** | **20 [6-103]** | **0.03** |
| Log Viral load | **<1.3 [<1.3-4.15]** | **5.37 [2.49-5.52]** | **0.0005** |
| **Active TB (% , n)** | **15.8 % (n=15)** | **13.2 % (n=5)** | 0.70 |
| **Co-morbidities** |  |  |  |
| Cardiovascular | **7.3% (n=7)** | **44.7% (n=17)** | **<0.0001** |
| Hypertension | **46.3% (n=44)** | **57.9% (n=22)** | 0.23 |
| Diabetes | **32.6% (n=31)** | **31.6% (n=12)** | 0.91 |
| Obesity | **26.3% (n=25)** | **34.2% (n=13)** | 0.36 |
| Other respiratory diseases | **2.1% (n=2)** | **28.9% (n=11)** | **<0.0001** |
| **WHO ordinal scale at enrolment (% , n)** |  |  |  |
| 2 | - | **13.2% (n=5)** | - |
| 3 | **17.9% (n=17)** | **44.7% (n=17)** | **0.0013** |
| 4 | **33.7% (n=32)** | **42.1% (n=16)** | 0.36 |
| 5 | **29.5% (n=28)** | - | - |
| 6 | **17.9% (n=17)** | - | - |
| 7 | **1% (n=1)** | - | - |
| **Mild and moderate (WHO <5, %)** | **51.6% (n=49)** | **100% (n=38)** | **<0.0001** |
| **Severe (WHO ≥5, %)** | **48.4% (n=46)** | - | - |
| **CRP (mg/L)** | **95 [42-152] (n=95)** | **25 [6-39] (n=35)** | **<0.0001** |
| **D-dimer (µg/mL)** | **0.68 [0.35-1.19] (n=90)** | **0.4 [0.1-0.61] (n=37)** | **0.0003** |
| **LDH (U/L)** | **409 [337-583] (n=93)** | **281 [222-410] (n=35)** | **<0.0001** |
| **Ferritin (ng/mL)** | **1123 [624-1846] (n=94)** | **131 [78-597] (n=35)** | **<0.0001** |
| **White cell count (x10<sup>9</sup>/L)** | **10.2 [7.1-13.5] (n=95)** | **10.3 [6.9-14.5] (n=33)** | 0.94 |
| **Unaffected lung (%)** | **30% [10-50] (n=86)** | **70% [50-80] (n=30)** | **<0.0001** |
| **On steroid treatment** | **78.9% (n=75)** | **28.9% (n=11)** | **<0.0001** |
| **Thrombo-embolic complications** | **10.5% (n=10)** | **0%** |  |
| **Days with symptoms prior sampling** | **9 [6-15]** | **8 [3-18]** | 0.49 |
| **Days in clinical care** | **15 [8-24]** | **6 [3-12]** | **<0.0001** |

29.5% (28/95) COVID-19 patients died and the comparisons of the clinical characteristics between discharged and deceased patients are presented in **Table S1**. As previously reported, COVID-19 patients who died were older, predominantly male, had more severe disease according to their WHO ordinal scale classification and were characterized by elevated systemic inflammation. No death occurred in the non-COVID-19 control group.

### Measures of COVID-19 severity

The WHO ordinal scale, stratifying patients according to their oxygen therapy requirement, has been widely used as correlate of COVID-19 severity. Additionally, a wide range of non-specific indicators of systemic inflammation (including amongst others: C-reactive protein (CRP), ferritin, serum amyloid A (SAA), procalcitonin, lactate dehydrogenase (LDH), D-dimer, IL-6, IL-10, white cell count or neutrophil count) have been associated with adverse COVID-19 outcome (Chen et al., 2020b; Izcovich et al., 2020; Szarpak et al., 2020; Zhang et al., 2020). Furthermore, higher levels of SARS-CoV-2-specific antibodies have been shown to associate with increased COVID-19 severity (Bosnjak et al., 2020; Klein et al., 2020). In this study cohort, we also observed increased level of SARS-CoV-2-specific antibodies in patients with severe COVID-19 defined by the WHO ordinal scale (**Figure S1**). Thus, based on the clinical data available in this study, eight clinical parameters were combined to perform a hierarchical clustering analysis, including WHO ordinal scale scoring, Roche Elecsys® anti-SARS-CoV-2 antibody cut-off index (COI), white cell count (WCC), CRP, D-dimer, Ferritin, LDH and radiographic evidence of disease expressed as % of unaffected lung. Two main clusters were identified: cluster 1 encompassed almost exclusively COVID-19 cases (92%), while cluster 2 contained 62.5% of hospitalized SARS-CoV-2 uninfected controls and 37.5% of COVID-19 cases. Moreover, two subgroups emerged from cluster 1, where cluster 1a was enriched in COVID-19 patients who died (**Figure 1A&B**). Principal component analysis (PCA) showed a good separation between COVID-19 cases and hospitalized non-COVID-19 controls with PC1 accounting for 32.7% and PC2 17.4% of the variance (**Figure 1C**). The corresponding loading plot shows that the lung % unaffected score, oxygen therapy requirement and white cell count were the main drivers of PC1 variance (**Figure 1D**). Furthermore, the PC1 score in COVID-19 patients who died was significantly higher (p < 0.0001) compared to patients who survived (**Figure 1E**). This analytical approach grades disease severity as a continuum, allowing the simultaneous integration of multiple clinical parameters of known relevance in COVID-19 outcome.

**Figure 1:**
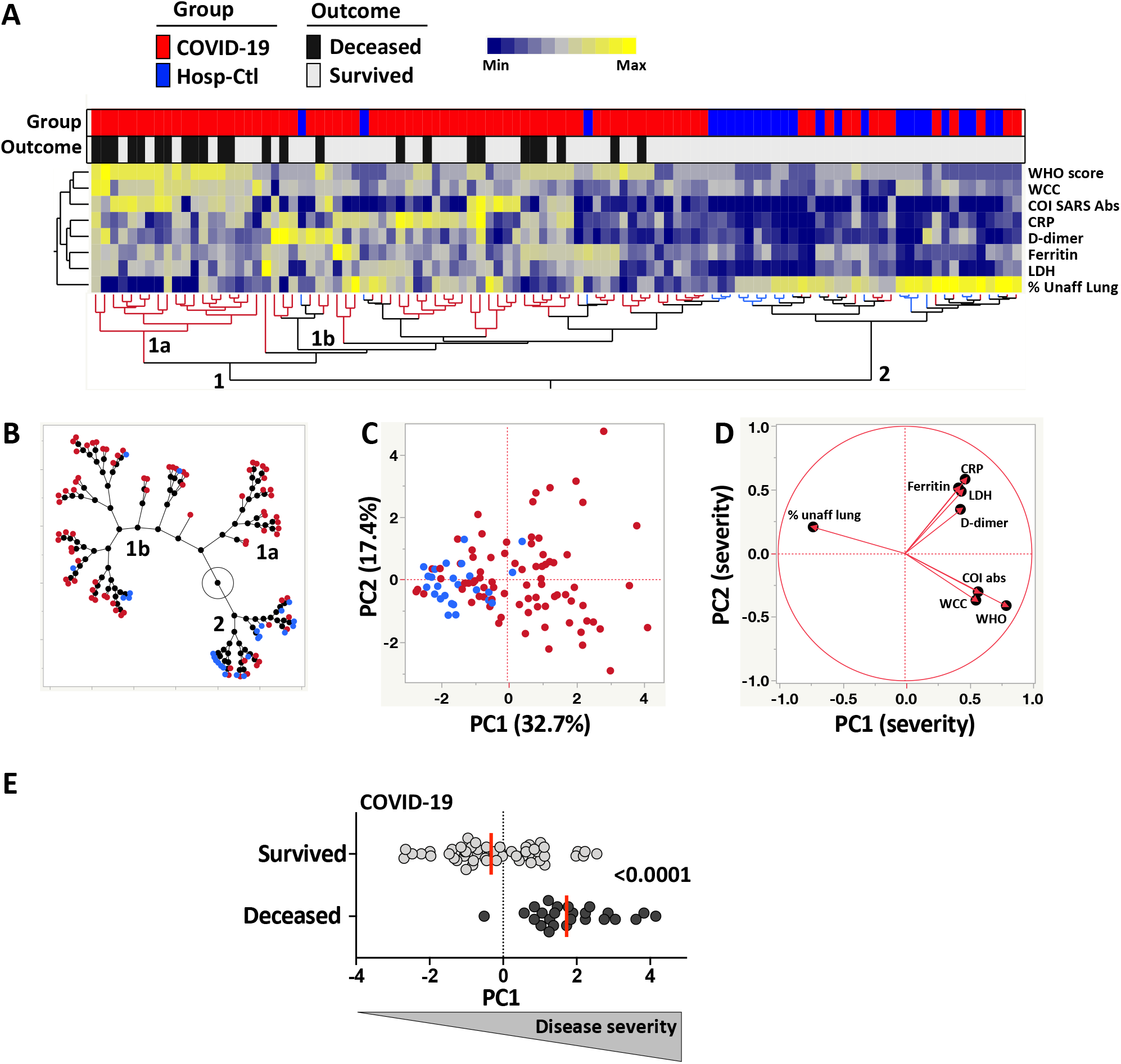
Measures of COVID-19 disease severity. **(A)** A non-supervised two-way hierarchical cluster analysis (HCA, Ward method) was employed to grade COVID-19 disease, using the WHO ordinal scale scoring, Roche Elecsys® anti-SARS-CoV-2 antibody cut-off index (COI), white cell count (WCC), C-reactive protein (CRP), D-dimer, Ferritin, lactate dehydrogenase (LDH) and radiographic evidence of disease extent expressed as % of unaffected lung. COVID-19 status (COVID-19 cases in red and SARS-CoV-2 uninfected hospitalized controls in blue) and outcome (survived in white and deceased in black) of each patient is indicated at the top of the dendrogram. Data are depicted as a heatmap colored from minimum to maximum values detected for each parameter. **(B)** Constellation Plot-cluster analysis based on all measured parameters. Each dot represents a participant and is color-coded according to their COVID-19 status. Each cluster obtained for the HCA is identified by a number. **(C)** Principal component analysis (PCA) on correlations, based on the eight clinical parameters, was used to explain the variance of the data distribution in the cohort. Each dot represents a participant. The two axes represent principal components 1 (PC1) and 2 (PC2). Their contribution to the total data variance is shown as a percentage. **(D)** Loading plot showing how each parameter influences PC1 and PC2 values. **(E)** Comparison of PC1 score values between COVID-19 cases who survived and those who died. Bars represent medians. Statistical comparisons were calculated using the non-parametric Mann-Whitney test.

### Distinct phenotype of SARS-CoV-2 responding CD4 T cells in COVID-19 and non-COVID-19 patients

First, we compared the prevalence, magnitude and phenotypic profile of SARS-CoV-2 responding CD4 T cells (e.g., cells producing IFNγ, TNFα or IL-2, **Figure 2A**) between hospitalized non-COVID-19 controls and confirmed COVID-19 patients. SARS-CoV-2 reactive CD4 T cells were detected in 34.2% (13/38) of non-COVID controls, while 83.2% (79/95) of COVID-19 patients exhibited a SARS-CoV-2-specific response (**Figure 2B**). This data concords with several publications demonstrating the presence of pre-existing SARS-CoV-2 cross-reactive CD4 T cells in 20 to 50% of SARS-CoV-2-unexposed individuals (Grifoni et al., 2020; Le Bert et al., 2020; Mateus et al., 2020; Rydyznski Moderbacher et al., 2020; Sekine et al., 2020; Weiskopf et al., 2020). We observed high variability in the magnitude of the SARS-CoV-2 CD4 T cell response amongst the SARS-CoV-2 responding participants from both groups; although not statistically significant, the median response in COVID-19 cases was ∼ 3-fold higher compared to non-COVID controls (0.17%, [IQR: 0.08-0.55] and 0.05%, [IQR: 0.03-0.36], respectively) (**Figure 2C**). Of note, in the COVID-19 group, the frequency of SARS-CoV-2 specific CD4 T cells strongly associated with the magnitude of SARS-CoV-2 nucleocapsid-specific IgG (*P* < 0.0001, r = 0.61) (**Figure S2A**), as previously reported (Demaret et al., 2020; Grifoni et al., 2020; Peng et al., 2020; Rydyznski Moderbacher et al., 2020).

**Figure 2:**
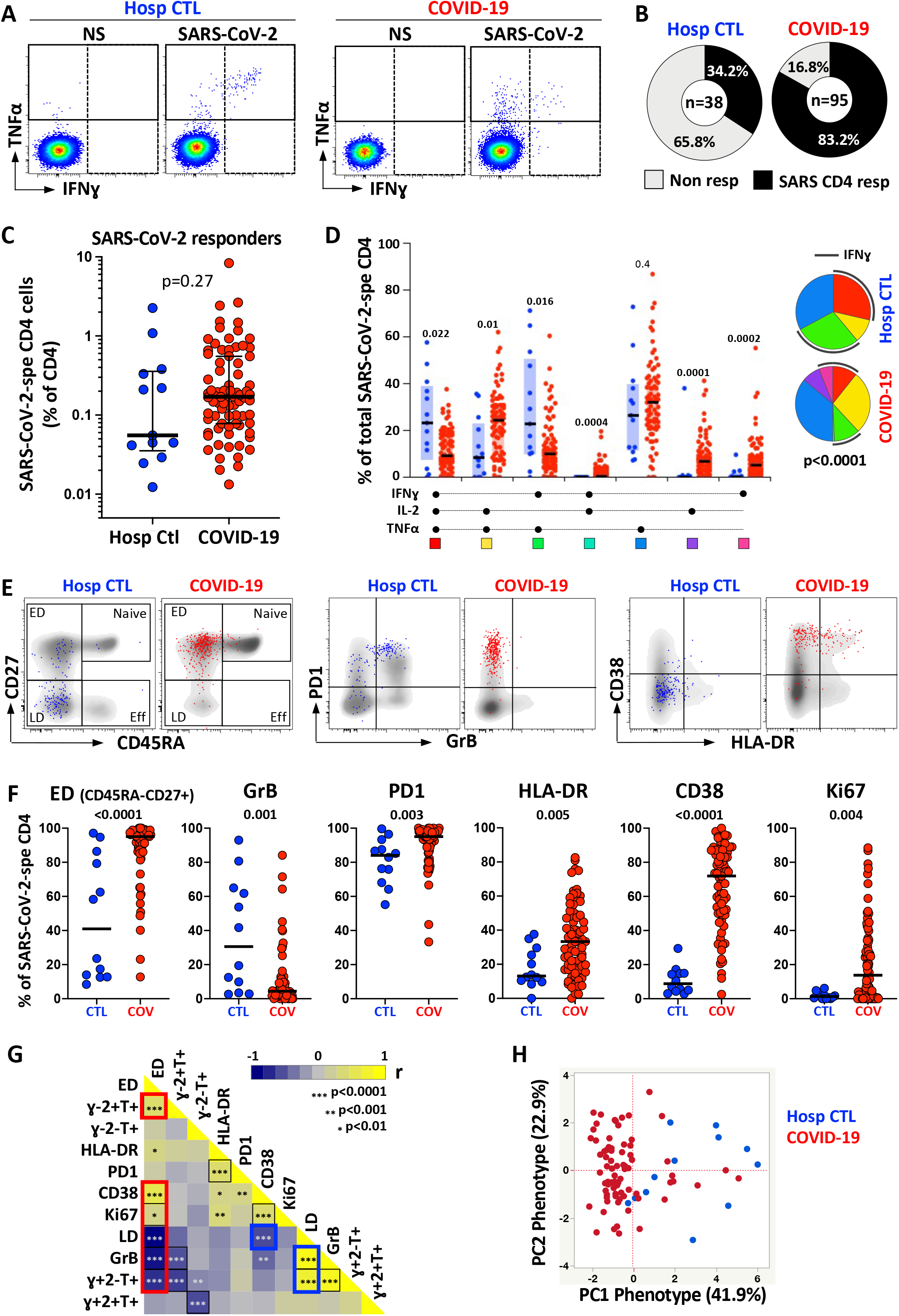
Prevalence, magnitude and functional profile of SARS-CoV-2-specific CD4 T cells between COVID-19 cases and SARS-CoV-2 uninfected hospitalized patients. **(A)** Representative flow cytometry plots showing the expression of IFNγ, and TNFα in SARS-CoV-2 -specific CD4 T cells in one SARS-CoV-2 uninfected control and one COVID-19 patient. NS = no stimulation; SARS-CoV-2 = stimulation with a pool of 15-mer peptides covering the M, N and immunogenic regions of the S proteins. **(B)** Proportion of patients exhibiting a detectable CD4 response to SARS-CoV-2 in the SARS-CoV-2 uninfected hospitalized control group and the COVID-19 group. The number of studied patients for each group is indicated at the center of the pie. **(C)** Comparison of the frequency of SARS-CoV-2 CD4 T cells in SARS-CoV-2 responders between SARS-CoV-2 uninfected hospitalized patients (blue) and COVID-19 cases (red). Statistical comparisons were performed using the Mann-Whitney test. **(D)** Comparison of the polyfunctional profile of SARS-CoV-2-specific CD4 T cells in hospitalized controls (blue) and COVID-19 cases (red). The x-axis displays each of the different response patterns, the composition of which is denoted with a dot for the presence of IFNγ, IL-2 and TNFα. The median (black bar) and interquartile ranges (box) are shown. Each response pattern is color-coded, and data are summarized in the pie charts, where every pie slice represents the median contribution of each response pattern to the total SARS-CoV-2 response. A Wilcoxon rank test was used to compare response patterns between groups (\*\*\**P* < 0.001, \*\**P* < 0.01, \**P* < 0.05). Statistical differences between pie charts were defined using a permutation test. **(E)** Representative overlay flow plots of CD45RA, CD27, PD1, GrB, CD38 and HLA-DR expression. Dots depict SARS-CoV-2-specific CD4 T cells in hospitalized controls (blue) and COVID-19 cases (red). Density plots in grey depict total CD4 T cells. Based on CD45RA and CD27 expression, four memory subsets can be delineated: Naive (CD45RA+CD27+), early differentiated (ED, CD45RA-CD27+), late differentiated (LD, CD45RA-CD27-) and effector (Eff, CD45RA+CD27-). **(F)** Summary graphs of the expression of each studied marker by SARS-CoV-2-specific CD4 T cells in both study groups. Bars represent medians. Statistical comparisons were performed using the Mann-Whitney test. **(G)** Heatmap of pairwise spearman correlations between phenotypic and functional traits of SARS-CoV-2-specific CD4 T cells. Spearman rank r correlation values are shown from blue (−1) to yellow (1). \*\*\**P* < 0.001, \*\**P* < 0.001, \**P* < 0.01. The red box identifies the profile of early differentiated SARS-CoV-2-specific CD4 T cells, predominantly observed in COVID-19 cases and the blue box the profile of late differentiated SARS-CoV-2-specific CD4 T cells enriched in SARS-CoV-2 uninfected hospitalized controls. **(H)** Principal component analysis (PCA) on correlations, based on the eight phenotypic and functional attributes of SARS-CoV-2-specific CD4 T cells (LD, GrB, HLA-DR, Ki67, CD38 and the proportion of IFNγ+IL-2-TNFα+ and IFNγ-IL-2-TNFα+). The two axes represent principal components 1 (PC1) and 2 (PC2). Their contribution to the total data variance is shown as a percentage.

When cytokine responses were analyzed individually, TNFα was the predominant cytokine produced by CD4 cells in response to SARS-CoV-2 peptides (using a short-term (5 h) whole blood assay) in both groups, with its production being significantly higher compared to IL-2 and IFNγ (**Figure S2B**). Combined analyses of all measured cytokines (IL-2, IFNγ and TNFα) showed that the overall polyfunctional profile of SARS-CoV-2-specific cells in COVID-19 participants was distinct from uninfected controls (*P* < 0.0001). In COVID-19, the CD4 response was characterized by limited expression of IFNγ and was enriched in cells co-expressing IL-2 and TNFα. Conversely, in non-COVID controls, most SARS-CoV-2-reactive CD4 T cells were distributed between triple functional cells (IL-2+IFNγ+TNFα+) and cells co-producing IFNγ and TNFα (**Figure 2D**).

We next assessed the memory differentiation (CD27, CD45RA), cytotoxic potential (GrB) and activation profile (HLA-DR, CD38, Ki67, PD-1) of SARS-CoV-2-responding CD4 T cells (**Figure 2E**). In COVID-19 patients, SARS-CoV-2-specific CD4 T cells almost exclusively displayed an early differentiated memory phenotype (ED: CD45RA-CD27+, median: 95.1%, [IQR: 88.7-97.4]). By contrast, in non-COVID controls, the memory profile of SARS-CoV-2-reactive CD4 T cells was highly variable between individuals, with 50% exhibiting predominantly a late differentiation profile (LD: CD45RA-CD27-). Moreover, the SARS-CoV-2 response in uninfected controls was characterized by significantly elevated expression of GrB compared to COVID-19 cases (median: 30.6%, [IQR: 5-64.2] vs 4.4%, [IQR: 1.9-9.6], respectively, *P* = 0.001). Interestingly, PD-1 was highly expressed on SARS-CoV-2-responding CD4 T cells in both clinical groups (median: 84%, [IQR: 91.8-99.6] in non-COVID controls and 95%, [IQR: 89.7-98.4] in COVID-19 patients), raising the question of potential intrinsic exhaustion of coronavirus-specific memory CD4 T cells (Jubel et al., 2020; Wherry and Kurachi, 2015; Wykes and Lewin, 2018).

As expected, the expression of HLA-DR, CD38 and Ki67 on SARS-CoV-2 responding CD4 T cells were significantly higher in COVID-19 cases compared to non-COVID-19 controls (*P* = 0.005, <0.0001 and 0.004, respectively), likely reflecting ongoing viral replication (**Figure 2F**). The expression of CD38 and Ki67 inversely associated with the time COVID-19 patients spent in clinical care (*P* = 0.0006, r = −0.39 and *P* = 0.017, r = −0.27, respectively, data not shown). As previously reported (Altmann and Boyton, 2020; Neidleman et al., 2020; Sekine et al., 2020), in convalescent COVID-19 patients (n=9), although the expression of HLA-DR, CD38 and Ki67 in SARS-CoV-2 CD4 T cells was significantly reduced compared to acute COVID-19 patients (reflecting viral clearance), cells maintained an elevated PD1 expression and retained their early differentiated phenotype (**Figure S2C**).

Pairwise associations of the functional and phenotypic characteristics of SARS-CoV-2 responding CD4 cells identified two signatures: 1) Activated cells exhibiting an early differentiated memory phenotype and preferentially secreting IL-2 and TNFα, characteristic of COVID-19 patients and 2) Late differentiated memory cells with elevated GrB expression endowed with polyfunctional capacities predominantly observed in SARS-CoV-2 responsive CD4 T cells from uninfected individuals (**Figure 2G**). To determine if the overall phenotypic profile of SARS-CoV-2-responding CD4 T cells allows discrimination between COVID-19-induced and pre-existing cross reactive CD4 responses, we performed a principal component analysis (PCA, **Figure 2H)** and hierarchical clustering analysis (**Figure S2D**), including eight parameters (e.g., the proportion of IFNγ+TNFα+IL2+,IFNγ-TNFα+IL2+ IFNγ-TNFα+IL2- and cells, the proportion of ED, and GrB, HLA-DR, CD38 and Ki67 expression). Both analyses showed that based on the functional and phenotypic traits of SARS-CoV-2 responding CD4 T cells, COVID-19 patients could be discriminated from non-COVID-19 controls.

### The functional and phenotypic signature of SARS-CoV-2-pecific CD4 T cells associates with disease severity

We next investigated the relationship between the profile of SARS-CoV-2-specific CD4 T cells and COVID-19 severity. While no difference was observed in the prevalence or magnitude of SARS-CoV-2-specific CD4 responses based on participant’s WHO ordinal scale score or outcome (survived vs deceased) (**Figure 3A**), their polyfunctional profile was related to disease severity. Less severe forms of disease were associated with enhanced capacity of SARS-CoV-2-specific CD4 T cells to co-express IFNγ, TNFα and IL-2. By contrast, TNFα monofunctional cells were more prevalent in patients with more severe disease (**Figure 3B**). These functional profiles also related to disease outcome (**Figure 3B**, inset). Assessing the phenotypic profile of SARS-CoV-2-specific CD4 T cells, the following trends were observed in less severe forms of COVID-19 (WHO ≤ 4): increased expression of CD38, Ki67 and GrB; and reduced expression of HLA-DR. However, PD1 expression and the memory maturation profile of SARS-CoV-2-specific CD4 T cells were comparable between COVID-19 patients, stratified by their WHO score. (**Figure 3C**).

**Figure 3:**
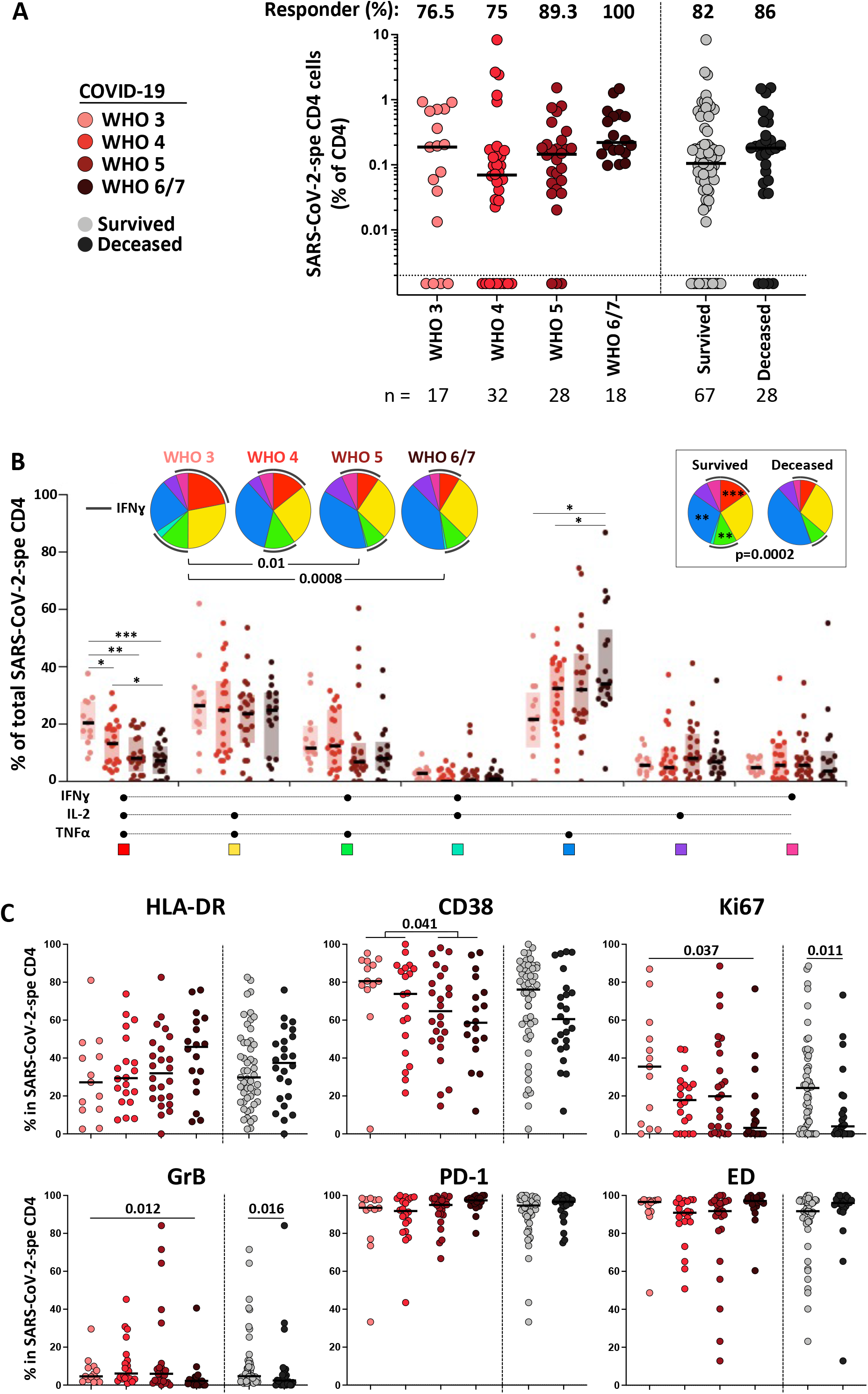
SARS-CoV-2-specific CD4 T cell response in COVID-19 cases stratified by WHO ordinal scale score and outcome. **(A)** Prevalence and frequency of SARS-CoV-2-specific CD4 T cells in COVID-19 cases. Patients were stratified according to WHO ordinal score and outcome. **(B)** Polyfunctional profile of SARS-CoV-2-specific CD4 T cells in COVID-19 cases, stratified by WHO score and outcome. The Wilcoxon rank test was used to compare response patterns between groups (\*\*\**P* < 0.001, \*\**P* < 0.01, \**P* < 0.05). Statistical differences between pie charts were defined using a permutation test. **(C)** Memory and activation profile of SARS-CoV-2-specific CD4 T cells in COVID-19 cases, stratified by WHO score and outcome. Statistical comparisons were defined using a Kruskal-Wallis test, adjusted for multiple comparisons (Dunn’s test) for the different WHO groups and the Mann-Whitney test to compare COVID-19 patients who survived or died.

Each functional and phenotypic attribute of SARS-CoV-2-specific CD4 T cells was assessed individually for the strength of its correlation with disease severity (defined by the composite analysis of clinical parameters described in Figure 1C (e.g., PC1 severity)). The highest Spearman rank r values for significant negative correlations were observed between the proportion of IFNγ+IL-2+TNFα+ cells, GrB and Ki67 expression and disease severity, while positive associations were found between the proportion of IFNγ-IL-2-TNFα+ cells, HLA-DR expression and disease severity (**Figure 4A**). Moreover, the global functional and phenotypic pattern of SARS-CoV-2-specific CD4 T cells described in Figure 2H (PC2 phenotype) associated with patients’ WHO ordinal scale score and outcome (survived vs deceased) (**Figure 4B**). Overall, COVID-19 severity (PC1 severity) strongly correlated with the traits of SARS-CoV-2-specific CD4 T cells (PC2 phenotype) (*P* = 0.0006, r = −0.43, **Figure 4C**), with severe disease being characterized by poor polyfunctional potential, reduced proliferation capacity and enhanced HLA-DR expression on SARS-CoV-2-specific CD4 T cells.

**Figure 4:**
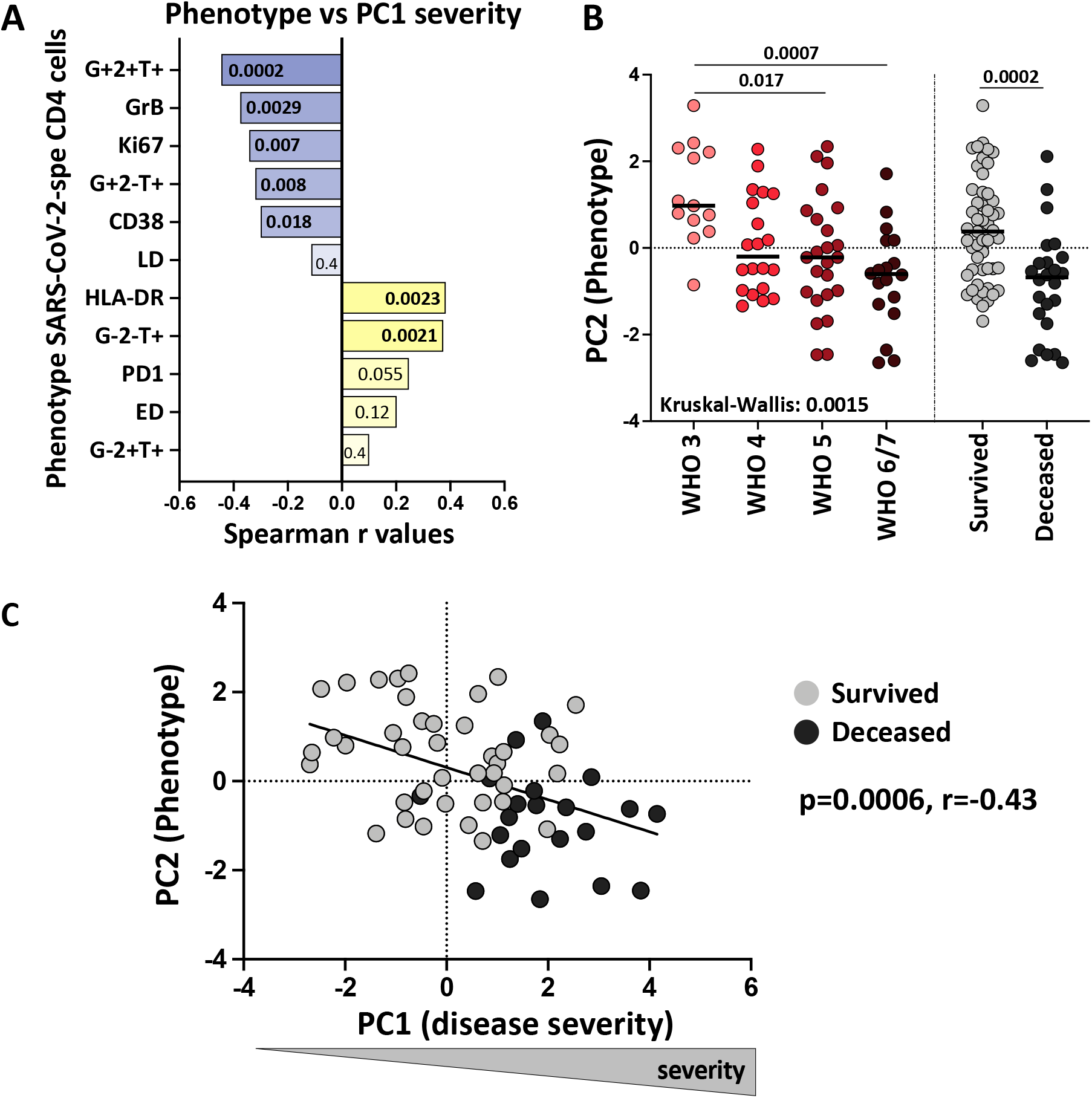
Relationship between COVID-19 severity and functional and phenotypical traits of SARS-CoV-2 - specific CD4 T cells. **(A)** Spearman Correlation r values between indicated SARS-CoV-2-specific CD4 T cell features and COVID-19 severity (defined by the composite analyze of clinical parameters, PC1 severity). Negative associations are represented in blue and positive associations in yellow. *P* values are indicated for each comparison. **(B)** Comparison of the overall profile of SARS-CoV-2-specific CD4 T cells (PC2 phenotype) in COVID-19 cases stratified by WHO ordinal score and outcome. Statistical comparisons were defined using a Kruskal-Wallis test, adjusted for multiple comparisons (Dunn’s test) for the different WHO groups and the Mann-Whitney test to compare COVID-19 patients who survived or died. **(C)** Association between COVID-19 severity (PC1 severity) and the overall profile of SARS-CoV-2-specific CD4 T cells (PC2 phenotype). COVID-19 survivors are depicted in grey and patients who died in black. Correlation was tested by a two-tailed non-parametric Spearman rank test.

### Pre-existing lymphopenia impairs the immune response to SARS-CoV-2 and current tuberculosis reduces the polyfunctional potential of SARS-CoV-2-specific CD4 T cells

Due to the systemic inflammation induced by chronic HIV infection and active TB, questions have been raised whether these two diseases in particular could distort the immune response to SARS-CoV-2, leading to increased mortality. Indeed, emerging evidence shows that TB and HIV are independently associated with an increased risk for COVID-19 mortality (Boulle et al., 2020; Geretti et al., 2020; Tamuzi et al., 2020). Thus, we defined the impact of HIV, TB and HIV/aTB co-infection on the magnitude, phenotypic and functional profile of the SARS-CoV-2 CD4 T cell response. Disease severity at enrolment (defined by PC1 severity or WHO ordinal scale on its own) was comparable irrespective of HIV and/or TB co-infection (**Figure 5A** and data not shown). However, age (an established risk factor for severe disease and mortality) could be a confounder as HIV+/aTB, aTB and HIV+ patients were significantly younger compared to the other COVID-19 patients (median: 40, 43, 47 and 55 years, respectively, **Figure S3A**).

**Figure 5:**
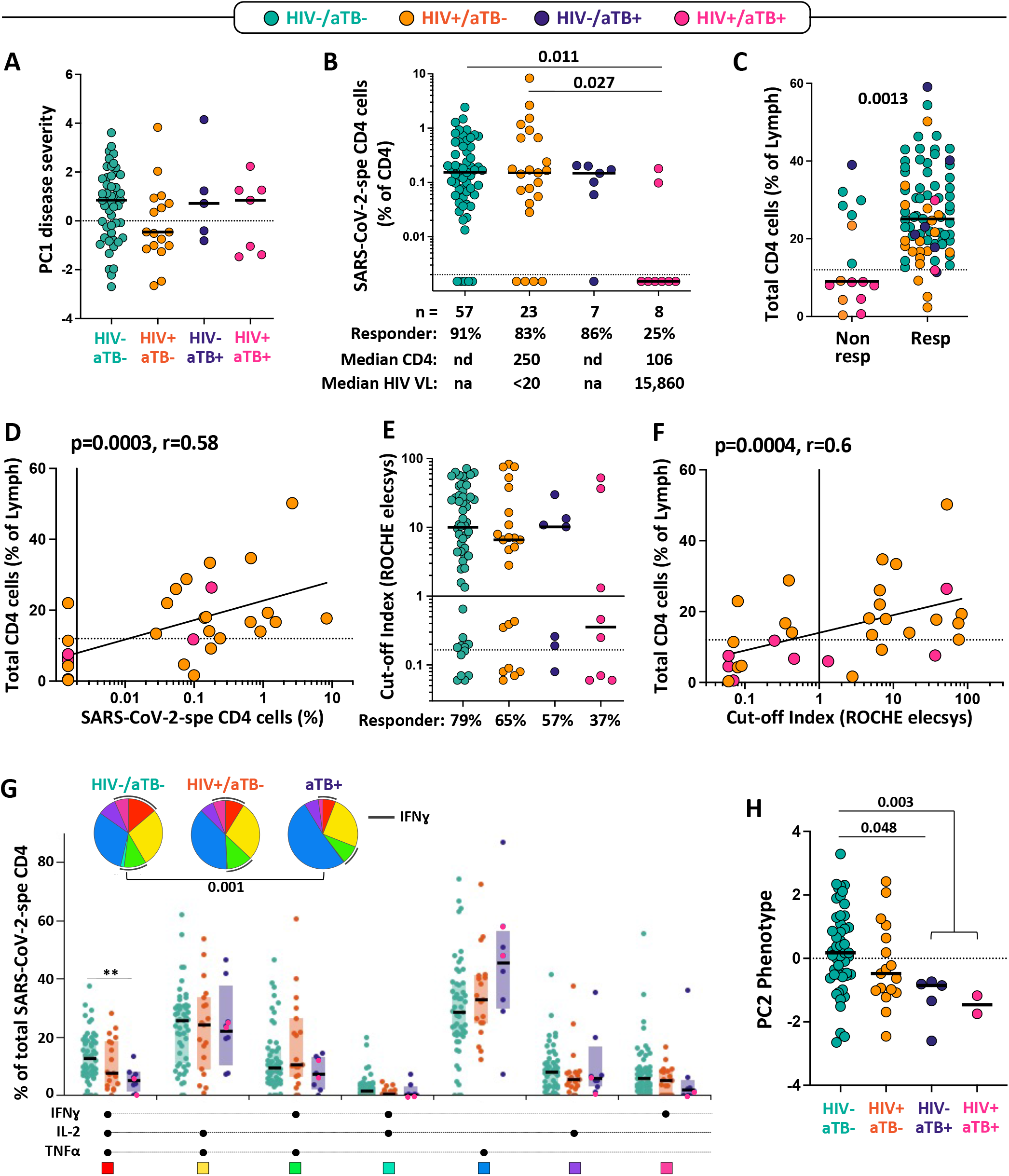
Impact of HIV, active TB (aTB) and HIV/aTB co-infection on SARS-CoV-2-specific CD4 T cell response. **(A)** Comparison of COVID-19 severity (defined by the composite analyze of clinical parameters, PC1 severity) between patients grouped according to HIV and/or aTB co-infection. **(B)** Prevalence and frequencies of SARS-CoV-2-specific CD4 T cells in COVID-19 patients stratified by HIV and/or aTB co-infection. Statistical comparisons were defined using a Kruskal-Wallis test, adjusted for multiple comparisons (Dunn’s test). **(C)** Comparison of the frequency of total CD4 T cells between SARS-CoV-2 CD4 responders and non-responders. Dots are color-coded according to patient’s HIV and TB status. Statistical comparison was performed using the Mann-Whitney test. **(D)** Association between the frequency of SARS-CoV-2-specific CD4 T cells and total CD4 T cells in HIV-infected COVID-19 patients. Correlation was tested by a two-tailed non-parametric Spearman rank test. **(E)** Prevalence and magnitude of SARS-CoV-2-specific serological response (defined using the Roche Elecsys® assay) in COVID-19 patients stratified by HIV and/or aTB co-infection. **F-** Association between the magnitude of SARS-CoV-2-specific serological response and the frequency of total CD4 T cells in HIV-infected COVID-19 patients. Correlation was tested by a two-tailed non-parametric Spearman rank test. **(G)** Polyfunctional profile of SARS-CoV-2-specific CD4 T cells in COVID-19 cases, stratified by HIV or aTB co-infection. For this analysis, HIV-/aTB+ and HIV+/aTB+ patients were combined in one group (aTB). Dots are color-coded according to patient’s HIV and TB status The Wilcoxon rank test was used to compare response patterns between groups (\*\**P* < 0.01). Statistical differences between pie charts were defined using a permutation test. **(H)** Comparison of the overall profile of SARS-CoV-2-specific CD4 T cells (PC2 phenotype) in COVID-19 cases stratified by HIV or aTB co-infection. Statistical comparisons were defined using a Kruskal-Wallis test, adjusted for multiple comparisons (Dunn’s test).

While the proportion of SARS-CoV-2 CD4 responders were similar between HIV-/aTB-, HIV+/ aTB- and HIV-/aTB+ patients (≥ 83%), in HIV-infected patients with aTB (HIV+/aTB+), only 25% (2/8) exhibited detectable SARS-CoV-2-specific CD4 T cells (**Figure 5B**). Of note, amongst responders, the frequency of SARS-CoV-2-specific CD4 T cells were comparable between all groups (**Figure 5B**). As HIV+/aTB+ patients are characterized by low absolute CD4 counts (median: 106 cells/mm^3^), we hypothesized that the lack of a SARS-CoV-2 specific response could be related to CD4 lymphopenia. As recent CD4 count data was not available for all patients, we used the frequency of total CD4 T cells, measured by flow cytometry, as a surrogate measurement of CD4 count, with the lowest frequencies of CD4 T cells being observed in participants with HIV-1+/aTB+ (**Figure S3B**). The frequency of total CD4 cells was significantly higher in SARS-CoV-2 responders compared to non-responders (median: 25% and 9%, respectively, *P* = 0.0013) (**Figure 5C**). Moreover, in HIV-infected patients, the magnitude of SARS-CoV-2-specific CD4 T cells associated with the frequency of total CD4 T cells (*P* = 0.0006, r = 0.58, **Figure 5D**) and absolute CD4 count (*P* = 0.001, r = 0.59, data not shown). Interestingly, patients co-infected with HIV and aTB also exhibited a limited capacity to generate SARS-CoV-2 antibodies with only 3 out of 8 patients having a positive serology of modest magnitude (**Figure 5E**). As for the frequency of SARS-CoV-2-specific CD4 response, the magnitude of SARS-CoV-2 antibodies correlated with the frequency of total CD4 T cells in HIV-infected patients (*P* = 0.0011, r = 0.56, **Figure 5F**). Of note, in our cohort, the lack of a SARS-CoV-2-specific CD4 response in patients with aTB was not associated with increased mortality, with death being recorded in 4 out of the 8 SARS-CoV-2 CD4 responders and 2 out of the 7 of CD4 non-responders (data not shown). We did not observe significant differences in the memory and activation profile of SARS-CoV-2-specific CD4 T cells based on patients’ HIV or TB status (**Figure S3C**). However, in COVID-19 patients with concomitant aTB, SARS-CoV-2-specific CD4 T cells displayed lower polyfunctional capacity, characterized by significant reduction of the cells with three functions, compared to HIV-/aTB-patients (**Figure 5G**). Finally, while HIV infection did not significantly alter the functional and phenotypic profile of SARS-CoV-2-specific CD4 T cell, in patients with aTB, the global SARS-CoV-2-specific CD4 T cell pattern was significantly different compared to HIV-uninfected COVID-19 patients (**Figure 5H**).

### Acute SARS-CoV-2 infection decreases *Mycobacterium tuberculosis* (Mtb)-specific CD4 T cell response

Many viruses, including SARS-CoV-2, cause a temporary immunosuppressive effect, which could lead to the reactivation of subclinical bacterial infection (Low et al., 2004). Thus, in a TB endemic country such as South Africa, many concerns have been raised about the possibility that COVID-19 could reactivate latent tuberculosis.

To better understand the potential impact of COVID-19 on Mtb co-infection, we compared the frequency and phenotype of Mtb-specific CD4 T cells in COVID-19 patients, hospitalized non-COVID controls and outpatient participants with latent TB (LTBI) or aTB recruited to unrelated studies prior to the emergence of the COVID-19 pandemic (**Table S2**). Mtb-specific T cell responses were also assessed using a whole blood assay (**Figure 6A**). As HIV infection is known to decrease Mtb-specific CD4 T cell response and aTB induces significant changes in the phenotype of Mtb-specific CD4 T cells (Riou et al., 2017), patients were grouped according to their HIV and TB status for this analysis. The proportion of CD4 responders to SARS-CoV-2 and Mtb were comparable in HIV-uninfected COVID-19 patients (∼90%). In HIV-infected patients with COVID-19, the proportion of a Mtb-specific CD4 response was significantly lower compared to that of SARS-CoV-2 (48% vs 83%, respectively, *P* = 0.013). Conversely, in COVID-19 patients with aTB, SARS-CoV-2 responses were only detected in 40% of participants, whilst 14/15 (93%) exhibited an Mtb-specific CD4 response (**Figure 6B**). We did not find any relationship between the extent of CD4 lymphopenia and the absence of a Mtb-specific responses (data not shown). Upon comparison of the frequency of Mtb-specific CD4 T cells between the current cohort and the 2018 pre-pandemic cohort with LTBI, we found that the magnitude of Mtb-specific CD4 T cells was ∼5-fold lower in the HIV-uninfected COVID-19 group and a ∼2-fold lower in the HIV-infected COVID-19 group compared to pre-pandemic samples (medians: 0.17% vs 0.53% for HIV-, *P* < 0.0001; and 0.09% vs 0.17% for HIV+, *P* = 0.052, respectively). However, comparable frequencies were observed in those with aTB (medians: 0.35% for COVID-19 vs 0.53% for pre-pandemic cohort, *P* = 0.3) (**Figure 6C**). This data suggests that acute SARS-CoV-2 infection may diminish the pool of Mtb-specific memory T cell responses.

**Figure 6:**
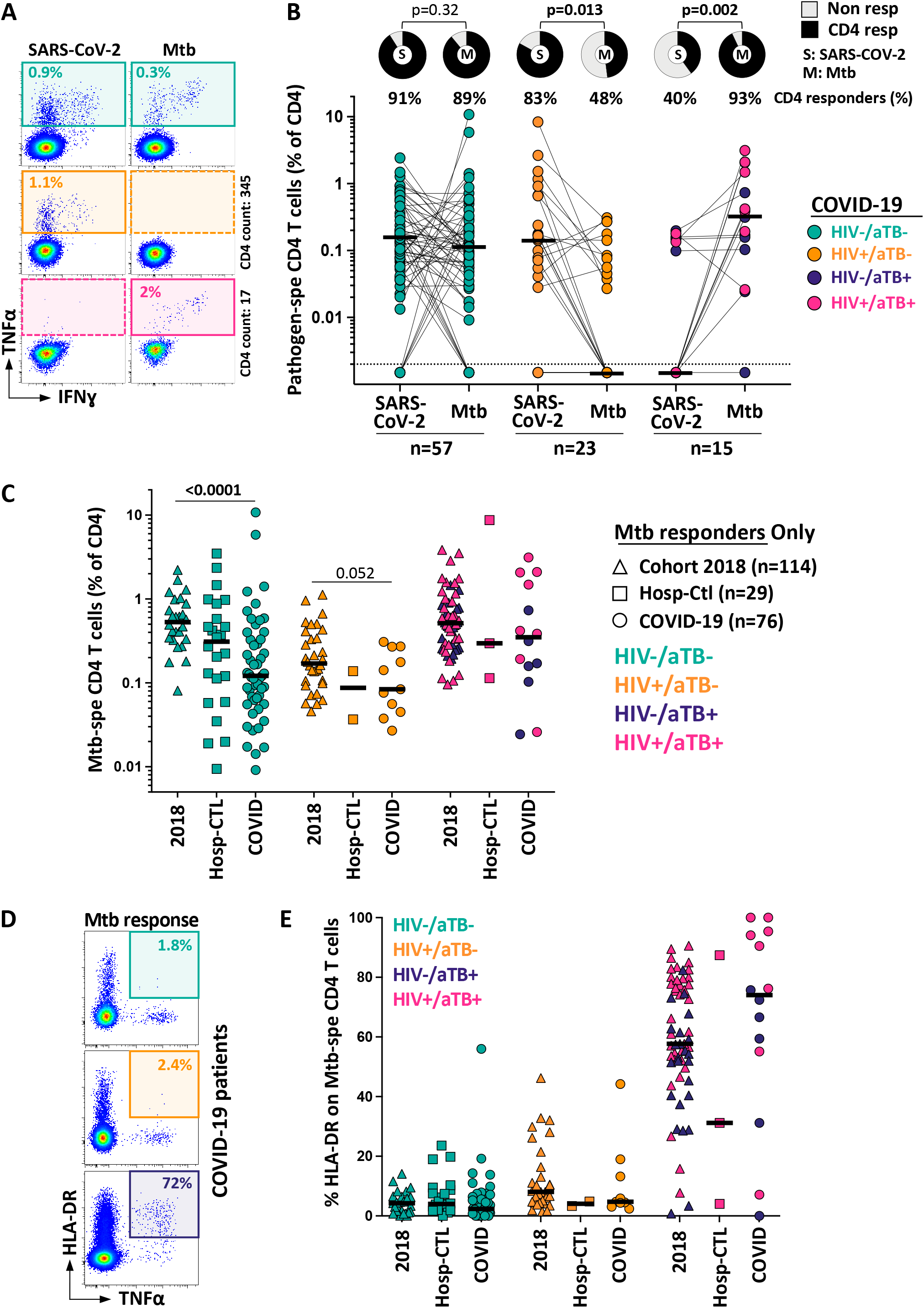
Impact of COVID-19 on Mtb-specific CD4 T cell response. **(A)** Representative examples of flow cytometry plots of SARS-CoV-2- and Mtb-specific CD4 T cell responses in three COVID-19 patients (one HIV-/aTB-, one HIV+/aTB- and one HIV+/aTB+). **(B)** Comparison of the prevalence and frequencies of SARS-CoV-2- and Mtb-specific CD4 T cells in COVID-19 patients stratified by HIV or aTB co-infection. The proportion of responders to each pathogen (S: SARS-CoV-2 and M: Mtb) is presented with pies at the top of the graph. Statistical comparisons were performed using the Chi-square test. Participants were grouped according to their HIV and/or TB status. Black bars represent the medians. **(C)** Comparisons of the frequencies of Mtb-specific CD4 T cells in a cohort recruited before the emergence of COVID-19 (2018, n = 114), SARS-CoV-2 uninfected hospitalized controls (n= 29) and COVID-19 cases (n = 76). Participants were stratified according to their HIV and/or TB status. Statistical comparisons were defined using a Kruskal-Wallis test, adjusted for multiple comparisons (Dunn’s test) for each sub-group. **(D)** Representative flow cytometry plots of HLA-DR expression on Mtb-specific CD4 T cells in three COVID-19 patients (one HIV-/aTB-, one HIV+/aTB- and one HIV+/aTB+). **(E)** Summary graph of HLA-DR expression on Mtb-specific CD4 T cells in a cohort recruited before the emergence of COVID-19 (2018), SARS-CoV-2 uninfected hospitalized controls and COVID-19 cases, stratified according to HIV and TB status.

Lastly, HLA-DR expression on Mtb-specific CD4 T cells has been shown to be a robust marker to discriminate active or subclinical TB from latent Mtb infection, regardless of HIV infection (Mpande et al., 2021; Riou et al., 2017). Thus, to define whether COVID-19 can promote Mtb reactivation, we compared the expression of HLA-DR on Mtb-specific CD4 T cells in the different cohorts (**Figure 6D**). In participants without aTB, no difference in the expression of HLA-DR was observed between COVID-19 patients, hospitalized non-COVID controls and the 2018 pre-pandemic cohort, irrespective of their HIV status (**Figure 6E**). Moreover, in these patients, the memory maturation profile and expression of other activation markers (such as CD38, Ki67 and PD1) in Mtb-specific CD4 T cells were similar between COVID-19 patients and hospitalized non-COVID controls (**Figure S4**). In aTB patients, elevated HLA-DR expression was observed compared to latently infected individuals, as expected. However, while not statistically significant, the proportion of activated Mtb-specific CD4 T cells tended to be higher in COVID-19 co-infected patients compared to the non-COVID-19 group (median: 74 % [IQR: 49-94] vs 57.7% [IQR: 50-77], respectively) (**Figure 6E**). This suggests that acute COVID-19 does not promote the reactivation of latent Mtb infection but could enhance the activation of the Mtb-specific CD4 T cell response during active TB.

## DISCUSSION

In this study, using a cohort of acute COVID-19 cases and SARS-CoV-2 uninfected hospitalized patients, we interrogated the SARS-CoV-2-specific CD4 T cell response patterns in relation to various measures of clinical disease severity to better understand the immune determinants of COVID-19 clinical course. Moreover, in a subset of patients, we investigated whether HIV and/or tuberculosis co-infections affected the CD4 response against SARS-CoV-2, and conversely whether COVID-19 impacted on the Mtb-specific CD4 response.

First, by measuring the prevalence of SARS-CoV-2-specific CD4 responses, we showed that SARS-CoV-2 reactive CD4 T cells were detected in a substantial proportion of SARS-CoV-2 uninfected patients (∼34%). This is in accordance with several studies reporting that SARS-CoV-2 cross-reactive memory T cells are detectable in 20 to 50% of individuals with no prior exposure to SARS-CoV-2 (Braun et al., 2020; Grifoni et al., 2020; Le Bert et al., 2020; Mateus et al., 2020; Rydyznski Moderbacher et al., 2020; Sekine et al., 2020; Sette and Crotty, 2020; Weiskopf et al., 2020). Limited information is available regarding the phenotype and function of these memory responses. Our data show that SARS-CoV-2-responding CD4 T cells were qualitatively different in acute COVID-19 cases compared to uninfected individuals. In the former group, SARS-CoV-2-specific CD4 T cells almost exclusively displayed an early differentiated memory phenotype and limited capacity to produce IFNγ; while in the latter group, SARS-CoV-2-responsive CD4 T cells preferentially exhibited a late differentiated memory phenotype and were enriched in GrB, suggesting that cytotoxic memory CD4 T cells could be a relevant component in SARS-CoV-2 immunity, as previously described for other viral infections (Juno et al., 2017). However, to date, the functional role for pre-existing cross-reactive T cell memory in COVID-19 remains unproven. In a comprehensive review, Leipsitch *et al*. describe three possible scenarios outlining potential mechanisms by which cross-reactive memory T cells could confer some form of protection against COVID-19, by reducing the viral burden and/or limiting disease severity or its duration (Lipsitch et al., 2020).

Several publications have reported that severe COVID-19 elicits drastic changes in the overall distribution and phenotypic landscape of circulating T cells, characterized by severe lymphopenia (preferentially affecting CD8 T cells) and widespread T cell activation (Chen and John Wherry, 2020; Laing et al., 2020; Mathew et al., 2020). Furthermore, an immune signature of the SARS-CoV-2-specific T cell response correlating with COVID-19 severity is also emerging (Meckiff et al., 2020; Oja et al., 2020; Rydyznski Moderbacher et al., 2020; Sattler et al., 2020; Schub et al., 2020; Weiskopf et al., 2020). Most of these studies compared patients with very divergent forms of disease (hospitalized vs non-hospitalized patients, convalescent patients who had mild or severe disease, or hospitalized vs convalescent patients). Here, we report on the immune profile of SARS-CoV-2 CD4 response in hospitalized acute COVID-19 patients stratified by disease severity based on multiple clinical parameters of known relevance in COVID-19 outcome. Our data show that the quality rather than the quantity of SARS-CoV-2-specific CD4 T cells may contribute to an efficient COVID-19 immune response as previously described for other viral infections (Seder et al., 2008). Indeed, more severe forms of COVID-19 correlate with the SARS-CoV-2-specific CD4 response displaying a limited capacity to produce IFNγ, reduced expression of GrB and Ki-67, and elevated expression of HLA-DR. This accords with other reports also showing that reduced IFNγ production characterizes severely ill patients (Oja et al., 2020; Schub et al., 2020). Moreover, the overall profile of SARS-CoV-2-specific CD4 T cells differed significantly between COVID-19 survivors and patients who died. The altered Th1 profile observed in severe COVID-19, reminiscent of an exhausted phenotype, could contribute to increased inflammation with poorer viral control. It thus remains to be seen whether recovery from COVID-19 would induce long lasting, efficient memory T cells, regardless of the severity of the COVID-19 episode.

In this study, we also report, for the first time, the impact of HIV, TB and HIV/TB co-infection on SARS-CoV-2 immunity. The clinical and epidemiological interactions of COVID-19 with tuberculosis and/or HIV-1 pose an additional health threat. In South Africa, two large epidemiological studies have shown that TB and HIV-1 were independently associated with increased risk of severe COVID-19 and death (Boulle et al., 2020; Davies, 2020). Although co-morbidities associated with HIV-1 and TB may primarily drive COVID-19 severity in these populations, it is also plausible that HIV and/or TB-associated immune dysregulation may contribute to heightened risk.

To date, the immunological impact of HIV on SARS-CoV-2 immune response has been mainly reported in isolated or limited cases of co-infection (d’Ettorre et al., 2020; Mondi et al., 2020; Wang et al., 2020). Only two studies have measured the effect of HIV-1 infection on the overall profile of T cells in COVID-19 cases: Karim *et al*. showed that viraemic HIV-infected COVID-19 patients exhibited lower frequencies of tissue homing CXCR3+ CD8 T cells and higher T cell activation compared to HIV-uninfected patients (Karim et al., 2020). Similarly, Sharov showed that viraemic HIV-infected COVID-19 patients displayed enhanced exhaustion of their T cell compartment (Sharov, 2020). This suggest that systemic immune activation associated with untreated HIV could skew the SARS-CoV-2 immune response. In our study, where most of the HIV+/aTB-participants were virally suppressed (17 out of 22), we showed that HIV infection alone did not alter the functional and phenotypic profile of SARS-Cov-2 CD4 T cells compared to HIV-uninfected patients. However, HIV-1 infected patients characteristically displayed a lower CD4 T cell frequency compared to HIV-uninfected patients, which in turn was associated with lower magnitudes of SARS-CoV-2-specific CD4 T cells and lower levels of IgG targeting SARS-CoV-2 nucleocapsid associated with total CD4 T cell frequency. Moreover, in most HIV+/aTB+ patients with the most severe lymphopenia (CD4 frequency < 10%), SARS-CoV-2-specific responses were undetectable. These results suggest that pre-existing lymphopenia, observed in untreated HIV-1 infection or in those with CD4 reconstitution despite ART usage, may impede the generation of T cell and/or antibody responses against SARS-CoV-2.

Immunological data on the SARS-CoV-2 response in the context of active TB co-infection is even more scarce (Gupta et al., 2020a). As both diseases can elicit a hyper-inflammatory state in the lung with overlap in the cytokine and chemokine profile found in broncho-alveolar lavage samples during severe COVID-19, TB or HIV/TB co-infection (Esmail et al., 2018; Liao et al., 2020), it can be speculated that one disease may exacerbate the other, leading to unfavorable outcomes. Here, we show that active TB co-infection skewed the functional profile of SARS-CoV-2-specific CD4 T cells, leading to a reduction of their polyfunctional capacity. It is possible that the excessive inflammation triggered by COVID-19 and TB co-infection underlies the premature functional exhaustion of SARS-CoV-2-specific T cells. Future studies to specifically examine differences in the inflammatory environment between COVID-19 and COVID-19/aTB patients in blood and in the lung would shed more light on the interplay between the two diseases.

Additionally, in countries where the prevalence of latent Mtb infection is high, the profound lymphopenia induced by SARS-CoV-2 and use of steroids as a treatment for COVID-19 could: 1) predispose patients to TB reactivation as a consequence of a transient suppression of cellular immunity and/or 2) increase the risk of progressive primary TB infection by reducing the pool of memory T cells targeting Mtb. We showed in this study that COVID-19 did not induce a concomitant activation of Mtb-specific CD4 T cells, suggesting that acute SARS-CoV-2 infection may not immediately result in progression of latent Mtb to subclinical or active TB disease. However, we found a significant reduction in the frequency of Mtb-specific CD4 T cells in COVID-19 patients compared to healthy pre-pandemic participants with LTBI. As an intact T cell response is an essential component in Mtb control, this decline in Mtb-specific CD4 T cells could impact the ability of the host to control latent or new Mtb infection. However, longitudinal studies are required to investigate whether T cell normalization after COVID-19 recovery is accompanied by homoeostatic re-expansion or peripheral redistribution of the Mtb-specific memory T cell pool. Furthermore, it remains to assess whether alterations in the frequency or phenotype of SARS-CoV-2 specific CD4 T cells, observed in the context of HIV and aTB co-infections, have an impact on COVID-19 clinical outcome, as the limited number of patients and the cross-sectional design of this study precluded speculation on this issue.

Overall, our results show that the functional and phenotypic signature of SARS-CoV-2-specific CD4 T cells, rather than magnitude, associates with COVID-19 severity in hospitalized patients. These results further advance our knowledge of the COVID-19 immunopathology, inform potential correlate of protection and could provide a rationale for future evaluation of novel vaccine responses. Moreover, our findings reveal potential mechanisms by which HIV-1 and TB co-infections could exacerbate COVID-19 pathology.

## Data Availability

This study did not generate any unique datasets or code.

## ACKNOWLEDGEMENTS

This work was supported by Wellcome Trust (104803, 203135 to RJW) and the National Institutes of Health (NIH) (R21AI148027 to CR). CR is supported by the European and Developing Countries Clinical Trials Partnership EDCTP2 programme supported by the European Union (EU)’s Horizon 2020 programme (Training and Mobility Action TMA2017SF-1951-TB-SPEC to CR). EdB is supported by a Harry Crossley Senior Clinical Fellowship.

The authors thank the study participants and their families, the clinical staff and personnel at Groote Schuur hospital in Cape Town for their support and dedication. We thank Dr Diana Hardie and Dr. Stephen Korsman at the Division of Medical Virology, National Health Laboratory Service, Groote Schuur Hospital for their assistance in obtaining the SARS-CoV-2 PCR cycle threshold values for the study participants. We thank Amanda Jackson and Celest Worship for the management of the clinical data. We also wish to thank Sheena Ruzive, Francisco Lakay, Nonzwakazi Bangani and Kennedy Zvinairo for their work on this project at the Wellcome Centre for Infectious Disease Research in Africa Laboratory at the University of Cape Town.

## AUTHOR CONTRIBUTIONS

CR, EdB, CS, RD and RJW designed the study. EdB, CS, RD and RTG recruited the study participants. RJW and SW facilitated clinical recruitment. EdB and CR performed the flow experiments. CR performed the data analysis and interpretation. FA supervised sample collection and processing. BWA conceptualized the radiographic reading method. QSH interpreted the radiological data. MH supervised the serology test. KAW, CSLA and AS provided critical reagents. CR wrote the manuscript with all authors contributing by providing critical feedback.

## DECLARATION OF INTEREST

The authors declare no conflict of interest.

## STAR METHODS

## RESOURCE AVAILABILITY

### Lead Contact

Further information and requests for detailed protocols should be directed to the lead contact, Dr. Catherine Riou.

### Materials Availability

With the exception of the Mtb peptide pool, all reagents used in this study are commercially available. The Mtb peptide pool was provided by Dr. Alessandro Sette and request regarding this peptide pool should be directed to Dr Sette.

### Data and Code Availability

The published article includes all data generated or analyzed during this study, and summarized in the accompanying tables, figures and supplemental materials.

## EXPERIMENTAL MODEL AND SUBJECT DETAILS

### Hospitalized COVID-19 and non-COVID-19 patients

We enrolled 133 hospitalized patients (95 with confirmed acute COVID-19 and 38 SARS-CoV-2 uninfected) from Groote Schuur Hospital in Cape Town, South Africa between June and August 2020. The clinical characteristics of all patients included in this study are presented in **Table 1** and the comparisons of the clinical characteristics between discharged and deceased COVID-19 patients are presented in **Table S1**. The University of Cape Town’s Faculty of Health Sciences Human Research Ethics Committee approved the study (HREC: 207/2020) and written informed consent was obtained from all participants with the capacity to provide it. Relatives provided proxy consent for those participants without capacity to consent for themselves (e.g., due to decreased level of consciousness). In cases where participants regained capacity, informed consent was obtained from them directly at that time.

### 2018 case-control study

To compare the frequency and profile of Mtb-specific CD4 T cells between samples collected before and during the SARS-CoV-2 pandemic, we used data generated from participants recruited at the Ubuntu Clinic, Site B, Khayelitsha (Cape Town, South Africa) between March 2017 and December 2018. This cohort has been described in detail in (Riou et al., 2020a) and the clinical characteristics of the study participants are shown in **Table S2**. Briefly, 122 adults (age ≥ 25), classified into four groups according to their HIV-1 and TB status: latent TB infection (LTBI) /HIV-(*n* = 24), LTBI/HIV+ (*n* = 30), active TB (aTB)/HIV- (*n* = 32), and aTB/HIV+ (*n* = 36), were included in this study. Median age was comparable between the four groups (median: 35 years [IQR: 31-45]). All active TB cases were sputum Xpert MTB/RIF (Xpert, Cepheid, Sunnyvale, CA) positive and had clinical symptoms and/or radiographic evidence of tuberculosis. The latent TB group were all asymptomatic, had a positive IFN-γ release assay (IGRA, QuantiFERON®-TB Gold In-Tube), tested sputum Xpert MTB/RIF negative and exhibited no clinical evidence of active TB. HIV-infected participants with LTBI had a significantly lower plasma HIV-1 viral load (VL) and higher absolute CD4 count compared to the HIV-infected aTB group. These differences are due to higher antiretroviral therapy (ART) usage in the LTBI group compared to the aTB group. All participants were provided written informed consent and the study was approved by the University of Cape Town Human Research Ethics Committee (HREC 050/2015) and was conducted under DMID protocol no.15-0047.

### Convalescent COVID-19 donors

Flow cytometry data were also available from a limited number of COVID-19 convalescent patients (n=9). These participants were healthcare workers recruited, between July and September 2020, from Groote Schuur Hospital in Cape Town. All had a SARS-CoV-2 PCR positive test, had mild symptoms, and did not require hospitalization. All participants were symptom-free at the time of sampling. Blood samples were obtained a median of 4.7 weeks post SARS-CoV-2 PCR test. The University of Cape Town’s Faculty of Health Sciences Human Research Ethics Committee approved the study (HREC: 207/2020) and written informed consent was obtained from all participants

## METHOD DETAILS

### Clinical data

At enrolment, participants’ clinical status was assessed according to the WHO ordinal scale based on their requirements for oxygen and supportive therapy (World Health Organization, 2020). WHO 2: Ambulatory with limitation of activities, WHO 3: Hospitalized without requiring supplemental oxygen, WHO 4: Hospitalized with oxygen therapy by mask or nasal prongs, WHO 5: Hospitalized and requiring non-invasive ventilation or use of high-flow oxygen devices, WHO 6: Hospitalized and receiving invasive mechanical ventilation and WHO 7: Hospitalized and receiving invasive mechanical ventilation and additional organ support such as extracorporeal membrane oxygenation (ECMO). Absolute CD4 count (for HIV-infected patients) and white cell counts (WCC) were obtained from patients’ medical files from the date closest to research blood collection. C-reactive protein (CRP), Ferritin, D-dimer, Lactate dehydrogenase (LDH) and HIV-1 viral load were measured from blood collected at enrolment. All clinical tests were performed by the South African National Health Laboratory Services (NHLS). Posteroanterior chest radiographs were assessed for the total percentage of the lung fields unaffected by any visible pathology. Thus, in the COVID-19 group this score quantified the percentage of normal lung that was not visibly affected by known features of COVID-19 pneumonia on the radiograph. In those with TB, or other respiratory infections this score similarly quantified the percentage of normal lung, not visibly affected by the relevant pathology on the radiograph. Those with a normal chest radiograph would thus score 100%.

### Measurement of SARS-CoV-2 nucleocapsid-specific IgG in plasma

SARS-CoV-2 specific antibodies were assayed by the Roche Elecsys^®^ Anti-SARS-CoV-2 immunoassay (Roche Diagnostics, Basel, Switzerland). This semi-quantitative electro-chemiluminescent immunoassay measures SARS-CoV-2 nucleocapsid-specific IgG. The assay was performed by the NHLS and interpreted according to manufacturers’ instructions (Roche: V 1.0 2020-05). Results are reported as numeric values in form of a cut-off index (COI; signal sample/cut-off), where a COI < 1.0 corresponds to non-reactive plasma and COI ≥ 1.0 to reactive plasma. At 14 days post-SARS-CoV-2 PCR confirmation, the sensitivity and specificity of the Elecsys^®^ Anti-SARS-CoV-2 immunoassay is reported as 99.5% (95% CI, 97.0 to 100.0%) and 99.80% (95% CI, 99.69 to 99.88%), respectively (2020; Favresse et al., 2020; Muench et al., 2020).

### Whole blood-based T cell detection assay

Blood was collected in sodium heparin tubes and processed within 3 h of collection. The whole blood assay was adapted from the protocol described by Hanekom *et al*. (Hanekom et al., 2004). We adapted this assay to detect SARS-CoV-2 specific T cells using synthetic SARS-CoV-2 PepTivator peptides (Miltenyi Biotec, Surrey, UK), consisting of 15-mer sequences with 11 amino acid overlap covering the immunodominant parts of the spike (S) protein, and the complete sequence of the nucleocapsid (N) and membrane (M) proteins (Riou et al., 2020b). All peptides were combined in a single pool and used at a final concentration of 1 µg/ml. Briefly, 400 µl whole blood was stimulated with the SARS-CoV-2 S, N and M protein peptide pool or a pool of 300 Mtb-derived peptides (Mtb300, 2 µg/ml) (Lindestam Arlehamn et al., 2016) at 37°C for 5 hours in the presence of co-stimulatory antibodies against CD28 and CD49d (1µg/ml each; BD Biosciences, San Jose, CA, USA) and Brefeldin-A (10µg/ml, Sigma-Aldrich, St Louis, MO, USA). Unstimulated blood was incubated with co-stimulatory antibodies, Brefeldin-A and an equimolar amount of DMSO. Red blood cell lysis and white cell fixation was performed in a single step using a Transcription Factor Fixation buffer (eBioscience, San Diego, CA, USA) for 20 minutes. Cells were then cryopreserved in freezing media (50% fetal bovine serum, 40% RPMI and 10% dimethyl sulfoxide) and stored in liquid nitrogen until batched analysis.

### Cell staining and flow cytometry

Cell staining was performed on cryopreserved cells that were thawed, washed and permeabilized with a Transcription Factor perm/wash buffer (eBioscience). Cells were then stained at room temperature for 45 min with antibodies for CD3 BV650, CD4 BV785, CD8 BV510, CD19-BV750, CD45RA Alexa 488, CD27 PE-Cy5, CD38 APC, HLA-DR BV605, Ki67 PerCP-Cy5.5, PD-1 PE, Granzyme B (GrB) BV421, IFNγ BV711, TNFα PE-Cy7 and IL-2, PE/Dazzle 594, as detailed in **Table S3**. Samples were acquired on a BD LSR-II and analyzed using FlowJo (v9.9.6, FlowJo LCC, Ashland, OR, USA). A positive response was defined as any cytokine response that was at least twice the background of unstimulated cells. To define the phenotype of SARS-CoV-2-specific CD4 T cells, a cut-off of 20 events was used.

## BIOINFORMATIC AND STATISTICAL ANALYSIS

Graphical representations were performed in Prism (v9; GraphPad Software Inc, San Diego, CA, USA) and JMP (v14.0.0; SAS Institute, Cary, NC, USA). Statistical tests were performed in Prism. Non-parametric tests were used for all comparisons. The Kruskal-Wallis test with Dunn’s multiple comparison test was used for group comparisons and the Mann-Whitney and Wilcoxon matched pairs test for unmatched and paired samples, respectively.

## SUPPLEMENTARY MATERIALS

## SUPPLEMENTARY FIGURE LEGENDS

**Figure S1** (Related to Fig. 1). Magnitude of SARS-CoV-2-specific serological response (defined using the Roche Elecsys® assay) in COVID-19 patients stratified according to WHO ordinal score and outcome. Statistical comparisons were defined using a Kruskal-Wallis test, adjusted for multiple comparisons (Dunn’s test) for the different WHO groups and the Mann-Whitney test to compare COVID-19 patients who survived or died.

**Figure S2** (Related to Fig. 2). **(A)** Association between the magnitude of SARS-CoV-2-specific serological response (defined using the Roche Elecsys® assay) and the frequency of SARS-CoV-2 CD4 T cells in COVID-19 patients. Correlation was tested by a two-tailed non-parametric Spearman rank test. **(B)** Comparison of frequency of IFNγ-, TNFα-, and IL2-in producing SARS-CoV-2-reactive CD4 T cells in acute COVID-19 cases (red) and hospitalized SARS-CoV-2-uninfected patients (blue). Only SARS-CoV-2 responders are depicted. **(C)** Comparison of the memory (left) and activation (right) profile of SARS-CoV-2-specific CD4 T cells between acute COVID-19 cases (red) and convalescent patients (green). Statistical comparisons were calculated using the non-parametric Mann-Whitney test. **(D)** Non-supervised two-way hierarchical cluster analysis (Ward method) of eight functional or phenotypic attributes of SARS-CoV-2-specific CD4 T cells (the proportion of IFNγ+TNFα+IL2+, IFNγ-TNFα+IL2+ IFNγ-TNFα+IL2- and cells, the proportion of ED, and GrB, HLA-DR, CD38 and Ki67 expression). COVID-19 status and outcome for each patient is indicated at the top of the dendrogram.

**Figure S3** (related to Fig. 5). **(A)** Comparison of the age of COVID-19 patients based on their HIV and TB status. **(B)** Comparison of the frequency of total CD4 T cells based on patient’s’ HIV and TB status. Statistical comparisons were performed using a Kruskal-Wallis test, adjusted for multiple comparisons (Dunn’s test). **(C)** Memory and activation profile of SARS-CoV-2-specific CD4 T cells in COVID-19 cases, stratified by HIV and/or aTB co-infection. ED: Early differentiated (CD45RA-CD27+).

**Figure S4** (related to Fig. 6) Comparison of the memory differentiation (left) and activation (right) profile of SARS-CoV-2- and Mtb-specific CD4 T cells in hospitalized COVID-19 and non-COVID-19 patients. Statistical comparisons were performed using a Kruskal-Wallis test, adjusted for multiple comparisons (Dunn’s test).

## SUPPLEMENTARY TABLE LEGENDS

**Table S1** (related to Table 1) **Clinical characteristics of COVID-19 patients who survived or died**. ^*^: SARS-CoV-2 serology was performed using the Roche Elecsys assay, measuring SARS-CoV-2 nucleocapsid-specific antibodies. ^**^: SARS-CoV-2 polymerase chain reaction (PCR) was performed using the Allplex™ 2019-nCoV Assay manufactured by Seegene. Medians are reported and numbers in brackets correspond to interquartile range [IQR]. ART: antiretroviral treatment, COI: Cut-off index of Roche Elecsys assay, CRP: C-Reactive protein, LDH: lactate dehydrogenase.

**Table S2** (related to Fig. 6) **Clinical characteristics of 2018 case-control cohort**. ART: anti-retroviral treatment. nd: not done, na: not applicable.

**Table S3** (related to method section) **List of antibodies used in the flow cytometry panel**.

## Notes

### Competing Interest Statement

The authors have declared no competing interest.

### Author Declarations

University of Cape Town IRB

