## Supplemental data for "Profile of SARS-CoV-2-specific CD4 T cell response: Relationship with disease severity and impact of HIV-1 and active *Mycobacterium tuberculosis* co-infection"

### Supplementary Table 1

| | **SURVIVED**<br>**70.5%** (n=67) | **DECEASED**<br>**29.5%** (n=28) | ***P-values*** |
|---|---|---|---|
| **Age** (median, IQR) | **50** [42-56] | **55** [44-65] | **0.029** |
| **Male** (%, n) | **50.7%** (n=34) | **75%** (n=21) | **0.029** |
| **HIV positive** (%, n) | **37.3%** (n=25) | **21.4%** (n=6) | 0.13 |
| on ART | **72%** (n=18) | **83.3%** (n=5) | 0.39 |
| Time on ART (years) | **9.5** [6-12] | **10** [3.5-11] | 0.75 |
| CD4 count (cells/$mm^3$) | **144** [53-332] | **113** [45-270] | 0.71 |
| Log Viral load | **<1.3** [<1.3-3.75] | **2.38** [<1.3-4.91] | 0.42 |
| **Active TB** (%, n) | **13.4%** (n=9) | **21.4%** (n=6) | 0.33 |
| **Co-morbidities** | | | |
| Cardiovascular | **7.5%** (n=5) | **7.1%** (n=2) | 0.96 |
| Hypertension | **43.3%** (n=29) | **53.6%** (n=15) | 0.36 |
| Diabetes | **34.3%** (n=23) | **46.4%** (n=13) | 0.25 |
| Obesity | **29.8%** (n=20) | **35.7%** (n=10) | 0.57 |
| Other respiratory diseases | **10.4%** (n=7) | **-** | **-** |
| **SARS-CoV2 serology positive**[*] | **67.2%** (n=45) | **75%** (n=21) | 0.45 |
| COI (median, IQR) | **5.5** [0.25-23.3] | **15.5** [0.73-39.3] | 0.057 |
| **WHO scale at enrolment** | | | |
| 3 | **25.4%** (n=17) | **-** | - |
| 4 | **43.3%** (n=29) | **10.7%** (n=3) | **0.002** |
| 5 | **22.4%** (n=15) | **46.4%** (n=13) | **0.019** |
| 6 | **8.9 %** (n=6) | **39.2%** (n=11) | **0.0004** |
| 7 | **-** | **3.6%** (n=1) | - |
| **Severe (WHO ≥5)** | **31.3%** (n=21) | **89.3%** (n=25) | **<0.0001** |
| **Cycle threshold SARS PCR**[**] | **30.6** [24.8-33.9] | **29.1** [25.3-34.2] | 0.83 |
| **CRP (mg/L)** | **66** [31-136] (n=67) | **129** [58-222] (n=28) | **0.015** |
| **D-dimer (µg/mL)** | **0.54** [0.32-0.98] (n=63) | **0.85** [0.59-2.0] (n=27) | **0.015** |
| **LDH (U/L)** | **385** [313-513] (n=66) | **543** [387-689] (n=27) | **0.0009** |
| **Ferritin (ng/mL)** | **921** [512-1581] (n=96) | **1719** [1031-2279] (n=28) | **0.002** |
| **White cell count ($x10^9$/L)** | **8.9** [6.3-12.2] (n=67) | **11.9** [9.0-16.6] (n=28) | **0.005** |
| **Unaffected lung (%)** | **40%** [20-70] (n=59) | **20%** [0-40] (n=27) | **<0.0001** |
| **On steroid treatment** | **73.1%** (n=49) | **92.9%** (n=26) | **0.032** |
| **Days in hospital** | **11** [6-23] | **15** [7-22] | 0.54 |

### Supplementary Table 2

| | HIV-/aTB- | HIV+/aTB- | HIV-/aTB+ | HIV+/aTB+ |
|---|---|---|---|---|
| **N** | **24** | **30** | **32** | **36** |
| **Age** (median, IQR) | **33** [28-43] | **34** [32-41] | **34** [32-42] | **37** [32-45] |
| **Male** (%) | **58.3%** | **23.3%** | **50%** | **66.7%** |
| **CD4 count** (cells/$mm^3$)† | nd | **481** [358-700] | nd | **268** [141 - 400] |
| **Log Viral load** | na | **<1.3** [<1.3-4.18] | na | **4.59** [2.19-5.00] |
| **On ART** (%) | na | **80.6 %** | na | **38.9%** |

### Supplementary Table 3

| Markers | Fluorochrome | Clone | Company | Cat. Number | Role |
|---|---|---|---|---|---|
| **CD3** | BV650 | OKT3 | BioLegend | 317323 | Lineage |
| **CD4** | BV785 | OKT4 | BioLegend | 317428 | |
| **CD8** | BV510 | RPA-T8 | BioLegend | 301048 | |
| **CD19** | BV750 | HIB19 | BioLegend | 302262 | |
| **CD45RA** | Alexa 488 | HI100 | BioLegend | 304114 | Memory differentiation |
| **CD27** | PE-cy5 | 1A4CD27 | Beckman | 6607107 | |
| **CD38** | APC | HIT2 | BD Bioscience | 555462 | Activation |
| **HLA-DR** | BV605 | L243 | BioLegend | 307640 | |
| **Ki67** | PerCP-cy5.5 | B56 | BD Bioscience | 561284 | |
| **PD-1** | PE | EH12.2H7 | BioLegend | 329906 | |
| **GrB** | BV421 | GB11 | BD Bioscience | 563388 | Cytotoxic potential |
| **IFNɣ** | BV711 | 4S.B3 | BioLegend | 502540 | Cytokine production |
| **TNFα** | PE-cy7 | MAB11 | BioLegend | 502930 | |
| **IL-2** | PE/Dazzle™ 594 | MQ1-17H12 | BioLegend | 500344 | |

### Supplementary Fig. 1

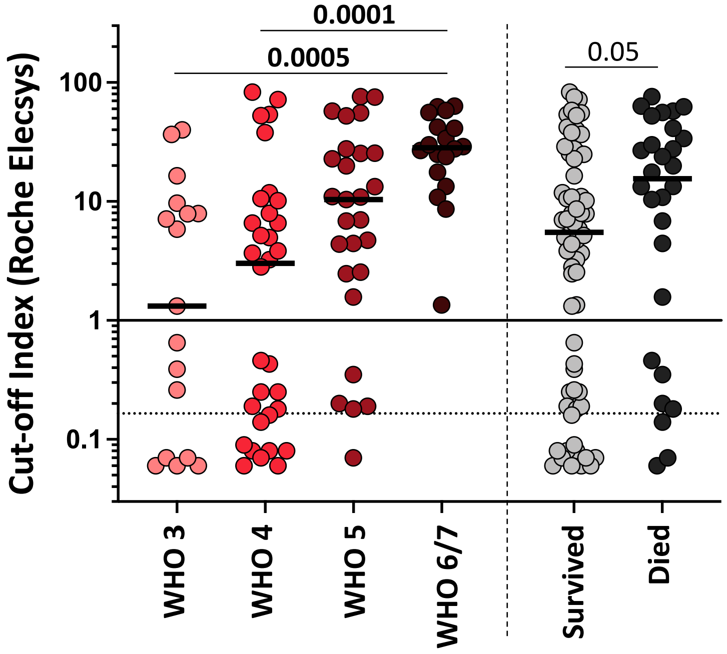

### Supplementary Fig. 2

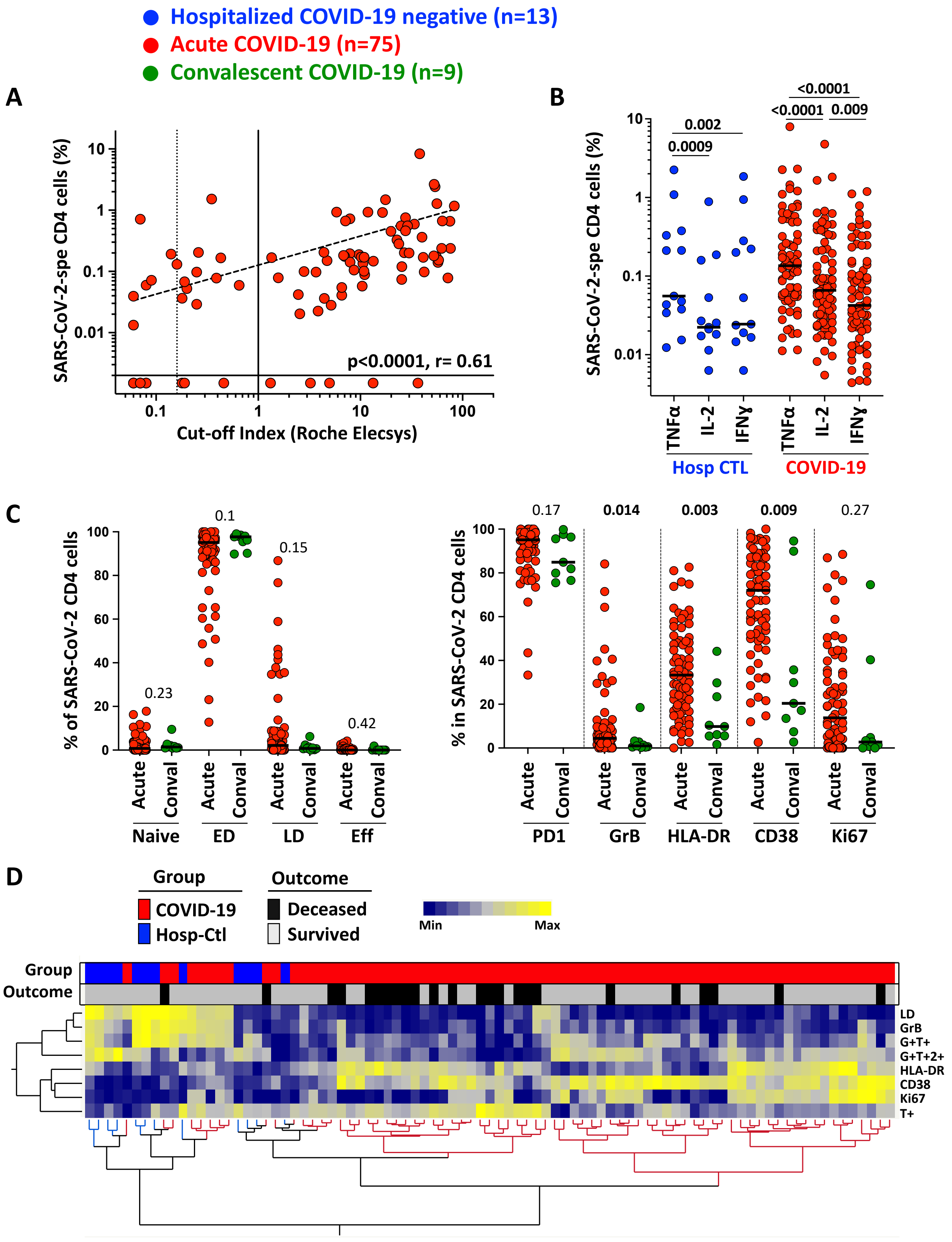

### Supplementary Fig. 3

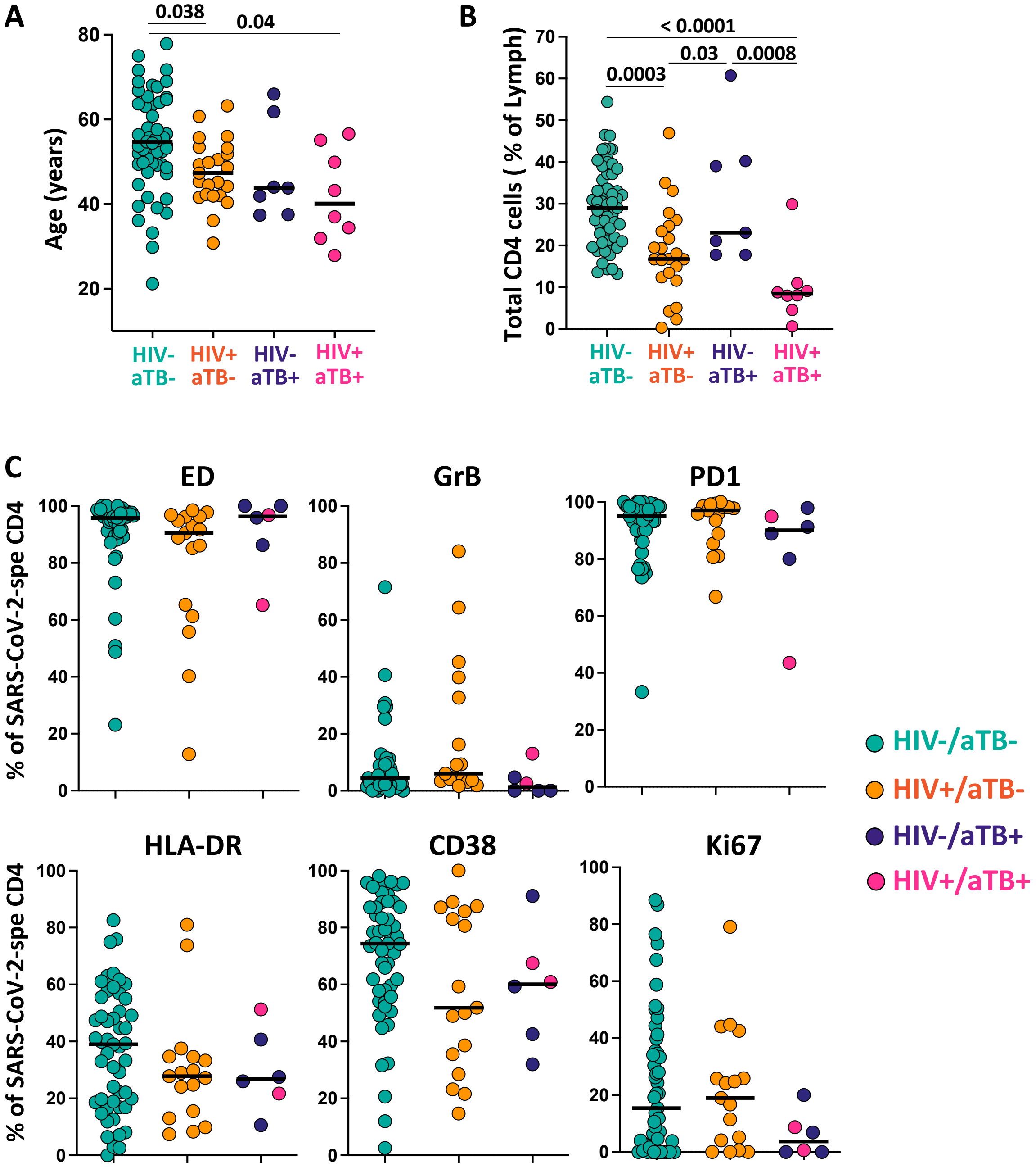

### Supplementary Fig. 4

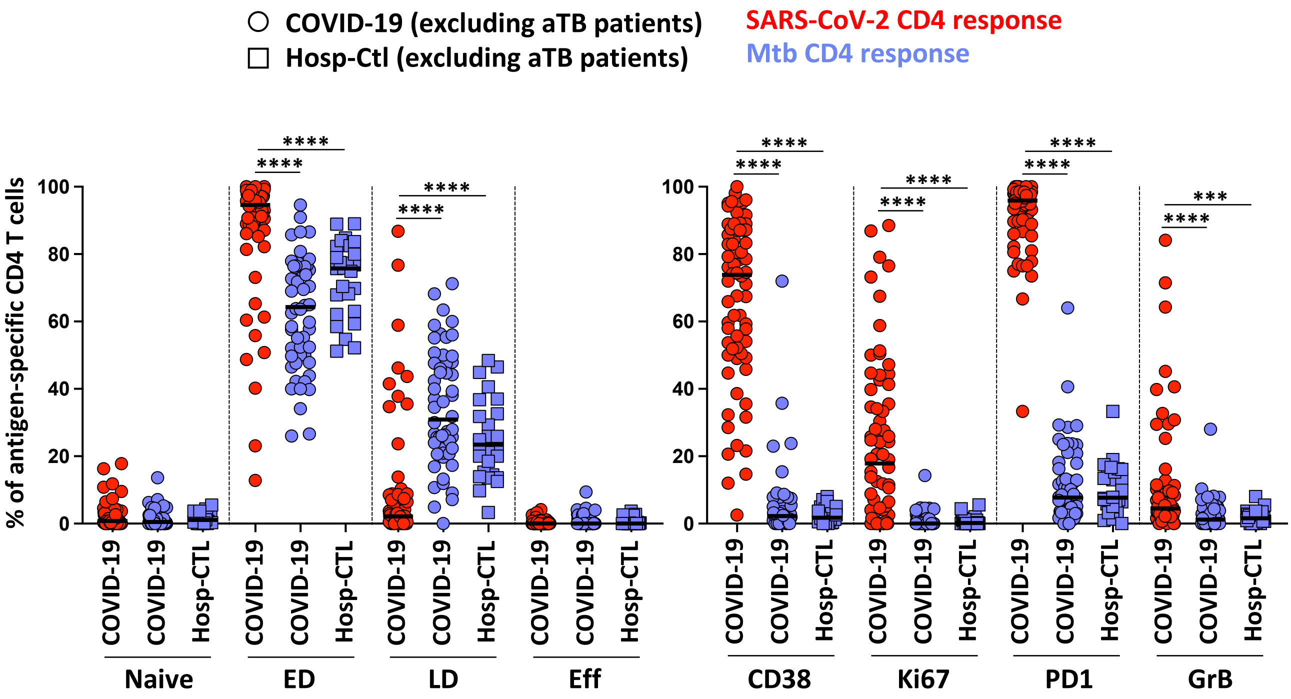
